# Modelling the role of human and vector behavioural patterns on the persistent transmission of *Plasmodium falciparum* malaria in Nigeria

**DOI:** 10.64898/2026.02.13.26346196

**Authors:** Idowu I. Olasupo, Emmanuel A. Bakare, Lukman O. Salaudeen

## Abstract

**Background:** Malaria, transmitted by female Anopheles mosquitoes, remains a major public health challenge in Nigeria, where approximately 97% of the population is at risk. Despite large-scale investments, Nigeria continues to bear the world’s highest malaria burden. Long-lasting insecticidal nets (LLINs) are central to prevention, yet their effectiveness is increasingly undermined by non-usage, delayed replacement, and growing outdoor biting activity. National surveys (MIS, PMI) consistently report gaps in LLIN use, irregular implementation of the three-year replacement strategy, and persistent outdoor biting. This study quantifies the relative contributions of these behavioural and entomological factors to sustained malaria transmission across five Nigerian states.

**Methods:** A deterministic compartmental model of malaria transmission was developed and calibrated using Bayesian inference with MCMC in CmdStanR. The model incorporated heterogeneous mosquito biting behaviour, LLIN effectiveness decay, and distribution cycles. Calibration used monthly malaria case data (2015-2024), demographic and entomological data (2015-2022), and DHS/MIS prevalence surveys (2015, 2018, 2021) for Akwa Ibom, Ebonyi, Kebbi, Oyo, and Plateau states. Counterfactual scenarios quantified malaria cases attributable to (i) outdoor mosquito biting, (ii) LLIN usage gaps, and (iii) delayed replacement. Parameter sweeps were used to assess how LLIN effectiveness changes with varying outdoor biting intensities.

**Results:** Eliminating outdoor biting yielded the largest reductions in malaria incidence—Akwa Ibom (82.4%, 95% CrI: 74.0-89.6), Ebonyi (92.0%, 95% CrI: 87.5-95.5), Kebbi (76.4%, 95% CrI: 51.0-92.6), and Oyo (83.0%, 95% CrI: 74.9-89.6). LLINs substantially reduced malaria transmission only under low outdoor biting intensities—below 1 bite per mosquito per month in Akwa Ibom and Ebonyi, below 0.2 in Kebbi and up to 0.8 in Oyo. In Plateau, outdoor biting contributed minimally (5.8%, 95% CrI: 4.8-6.3), while the gap between LLIN ownership and use was the dominant factor (23.1%, 95% CrI: 22.5-23.8), rising to 36.5% (95% CrI: 35.5-37.6) when combined with delayed replacement.

**Conclusion:** Outdoor mosquito biting is a dominant driver of persistent malaria transmission in Akwa Ibom, Ebonyi, Kebbi, and Oyo states, whereas low LLIN usage is the leading factor in Plateau. Although maintaining high LLIN coverage, adherence, and timely replacement remains critical, these efforts alone are insufficient where outdoor biting is widespread. Strengthening Nigeria’s malaria control strategy will require integrating LLIN deployment with targeted outdoor vector control and state-specific behavioural interventions to achieve sustained reductions in malaria burden.

## 1 Introduction

Malaria is known as a vector-borne disease caused by parasites of the *Plasmodium* family and transmitted by female *Anopheles* mosquitoes (WHO, 2015, 2022). It is of utmost public health concern because of its devastating impact on people’s health and livelihoods around the world (WHO, 2015). Five parasite species of *Plasmodium* have been documented to cause malaria disease in humans: *Plasmodium falciparum* (*P. falciparum*), *Plasmodium vivax* (*P. vivax*), *Plasmodium ovale* (*P. ovale*), *Plasmodium malariae* (*P. malariae*), *Plasmodium knowlesi* (*P. kwolesi*) (NMEP, 2021).

In 2023, there were an estimated 263 million malaria cases globally, implying an increase of 11 million cases from the previous year (WHO, 2024). Nigeria remains the country with the highest malaria burden in the world, accounting for 26% of global malaria cases, 30.9% of global malaria death and 39.3% of global malaria deaths in children aged under 5 years (WHO, 2024). An estimated 97% of Nigeria’s population are at risk of getting the disease (NMEP, 2021).

Efforts to curtail malaria menace had seen Nigeria and her partners commit significant human, financial and material resources to reduce the malaria burden and to work towards achieving a malaria-free status (NMEP, 2021). Every year, huge amount of money goes into financing various control interventions in Nigeria, ranging from vector control; drug-based prevention such as seasonal malaria chemoprevention (SMC) for children under-five years of age and intermittent preventive treatment in pregnancy (IPTp); case management involving diagnosis and use of artemisinin based combination therapy (ACT) for treatment (NMEP, 2021; PMI, 2024).

Vector control remains a vital component of malaria control and elimination strategies (WHO, 2012). Long-lasting insecticidal nets (LLINs) are the primary vector control intervention used in Nigeria (USAIDs, 2019; NMEP, 2021). The mode of operation of LLINs is that, they inhibit mosquito blood feeding by repelling mosquitoes and by posing a physical barrier to mosquitoes, and they kill adult female mosquitoes that attempt to bite a protected individual upon contact with insecticide in the nets (Yakob and Yan, 2009; Davis et al., 2020; NMEP, 2022). It is known that LLINs are expected to retain biological activity for at least 20 standard World Health Organization (WHO) washes under laboratory conditions and 3 years of recommended use under field conditions (WHO, 2005; Randriamaherijaona et al., 2017), however, there is usually variation across different settings and for different bed net types (Obi et al., 2020; Hiruy et al., 2023). A three-year durability monitoring study of LLINs (DAWA Plus 2.0, polyester treated with deltamethrin -distributed during the 2015/16 mass distribution campaign in Nigeria) in three locations with different ecological, demographic, and behavioural environments was conducted using a cohort of households representing the selected local government areas and followed up approximately 12, 24, and 36 months after distribution (Obi et al., 2020). The outcome of median net survival was estimated as the time in years until 50% of the originally distributed LLINs were no longer serviceable and it was found that median survival of LLINs is 5.3, 3.3 and 3.2 years in Zamfara, Ebonyi and Oyo states respectively (Obi et al., 2020). Also, the 24- and 36-month samples taken for insecticidal effectiveness testing (bio-assay) from all three sites showed a relatively low knockdown rate of 50% to 69%, but 24-hour mortality was above 95% for all samples, resulting in 100% optimal performance in Ebonyi and Oyo and 97% in Zamfara, showing optimal insecticidal performance based on WHO criteria (i.e ability of LLINs to cause mortality > 80% and or blood-feeding inhibition > 90% at the end of 3 years) (WHO, 2005; Obi et al., 2020).

Although LLINs are widely recognized as an effective vector control intervention for malaria, their effectiveness can be undermined by several factors (Savi, 2022). Studies have indicated that human behaviour can have a debilitating effect on the performance of LLINs in reducing malaria transmission, for instance, Ordinioha (2012); Mosha et al. (2020); Zemene et al. (2021) examined the extent to which distributed insecticide-treated nets (ITNs) are actually used in various countries and revealed a notable gap between ITN ownership and actual usage. In particular, the Malaria Indicator Survey (MIS) has also documented significant gaps between LLIN ownership and actual usage, as well as delays in the replacement of worn-out nets across various states in Nigeria (NMEP, 2022). These limitations, categorized as human behavioural factors, have the potential to reduce the protective impact of LLINs (Yakob and Yan, 2009; Savi, 2022). Additionally, LLINs are primarily designed for indoor use during sleeping hours; however, mosquito biting is not confined to indoor environments (Milali et al., 2017; Fernandez Montoya et al., 2022). Studies have reported exhibition of heterogeneous biting pattern by mosquitoes, probing into how residual early and outdoor biting can sustain malaria transmission in different African countries (Afolabi et al., 2006; Oyewole et al., 2007; Milali et al., 2017; Moshi et al., 2018; Sherrard-Smith et al., 2019; Irikannu et al., 2020; Degefa et al., 2021; Tomas et al., 2022; Khatib et al., 2025). Evidence from the President’s Malaria Initiative (PMI) indicates notable levels of outdoor mosquito biting activity across different Nigerian states (PMI, 2015, 2016, 2017, 2018, 2019, 2020, 2021, 2022). This outdoor biting behaviour poses another challenge to the effectiveness of LLINs, as it exposes individuals to mosquito bites beyond the protective coverage of the nets (Milali et al., 2017; Davis et al., 2020; Monroe et al., 2020; Fernandez Montoya et al., 2022; WHO, 2022).

Mathematical modelling has been used as a tool to investigate different aspects of vector control intervention in malaria transmission dynamics (Killeen et al., 2006; Agusto et al., 2013; Ngonghala et al., 2014; Bakare, 2015; Gimba and Bala, 2017; Collins and Duffy, 2022; Davis et al., 2024). Killeen et al. (2006) developed an analytical model which was applied to estimate the effective protection provided by an ITN, based on published experimental hut trials combined with questionnaire surveys of human sleeping behaviour and recorded mosquito biting patterns. In their study, they discovered that An. gambiae was predominantly endophagic and nocturnal in both surveys: Approximately 90% and 80% of exposure occurred indoors and during peak sleeping hours, respectively. ITNs consistently conferred > 70% protection against exposure to malaria transmission for users relative to non-users. Agusto et al. (2013) formulated and analyzed a mathematical model that looks at the transmission dynamics of malaria infection in mosquito and human populations and assessed the effects of bed-nets on its control. In their work, they found that if 75% of the population were to use bed-nets, malaria could be eliminated. Ngonghala et al. (2014) develop a mathematical model for malaria spread that captures the decrease in ITN effectiveness due to physical and chemical decay, as well as human behaviour as a function of time. For the case in which ITN efficacy decays over time, they determined coverage levels required to control malaria for different ITN efficacies and established that ITNs with longer useful lifespans have better performance in malaria control. Bakare (2015) formulated and analysed a mathematical model for malaria with multiple vector control. In their study, they found that in the presence of multiple vector control strategies the malaria disease transmission is reduced to a minimal level in the population. Gimba and Bala (2017) developed a compartmental deterministic model with incorporation of temperature and usage of insecticides treated bed nets to determine the impact of insecticide treated nets (ITNs) use, temperature, and treatment on malaria transmission dynamics. Their results show that the peak of mosquitoes biting rate occurs at a range of temperature values not on a single value as previously reported in literature and that the combination of treatment and ITNs usage is the most effective intervention strategy towards control and eradication of malaria transmissions. Collins and Duffy (2022) used a mathematical model fitted to data on the incidence of malaria in Nigeria to study the dynamics of malaria in Nigeria incorporating drug resistance, treatment, and the use of mosquito nets. Their results indicated that the disease is likely to remain endemic in Nigeria unless better control measures are focused on the dominant resistant strain, treatment is improved and the use of mosquito nets become widespread. Davis et al. (2024) analytically derived the steady-state solution of an age-structured deterministic compartmental model describing the mosquito gonotrophic cycle, with the age of each mosquito measured in the number of gonotrophic cycles (or successful blood meals) completed. Aside analytical derivations, they also investigated the impact of combinations of commonly used vector control methods on the age-structure of the vector population. They found that whilst well-maintain adult-acting vector control measures are substantially more effective than larval-based interventions, incorporating larval control in existing LLIN or IRS programmes could substantially reduce transmission.

While some studies have investigated malaria transmission dynamics in Nigeria, to the best of our knowledge, none has employed state-level historical malaria data to calibrate a mathematical model for quantifying the specific contributions of outdoor mosquito biting, the gap between LLIN ownership and usage, and delays in the replacement of worn-out LLINs. This study aims to retrospectively assess the impact of these factors on malaria incidence across selected Nigerian states, each representing distinct ecological zones. Hence, the question at the heart of this study is, how much of the reported malaria cases in the past ten years (2015 to 2024) is attributable to each of the identified human and vector behaviours in Nigeria? The overarching objective of this retrospective study is to determine the most influential drivers of sustained malaria transmission within each zone, thereby providing data-driven insights to inform and optimize vector control policy and intervention strategies in Nigeria.

The study will employ historical malaria data to calibrate a deterministic compartmental model, which will then be used to simulate a series of counterfactual scenarios. These simulations will estimate the potential reduction in malaria incidence under hypothetical conditions (counterfactual scenarios) where (i) there is no discrepancy between LLIN ownership and actual usage, (ii) the recommended three-year LLIN replacement interval is strictly followed, and (iii) outdoor mosquito biting is eliminated. The difference in malaria case counts between these counterfactual scenarios and the baseline situation will provide estimates of the number of cases attributable to each of the targeted human and vector-related behavioural factors in the ongoing transmission of malaria.

The remainder of this article is structured as follows: Section 2 outlines the materials and methods, encompassing the study area, data description, model formulation, and quantitative analysis. Section 3 presents the results and Section 4 discussion of the findings. Finally, Section 5 concludes with a summary of key insights, limitations of the study, and recommendations.

## 2 Materials and Methods

### 2.1 Study Areas

Nigeria, the most populous country in Africa, covers a land area of 923,708 sq. km (NMEP, 2021). Administratively, it consists of six geopolitical zones (North West, North East, North Central, South West, South South, and South East), 36 states plus the Federal Capital Territory (Abuja), and 774 local government areas (LGAs) (NMEP, 2021; USAIDs, 2019). Malaria transmission intensity and seasonality vary across five ecological zones—mangrove swamp, rain-forest, Guinea savannah, Sudan savannah, and Sahel savannah—each shaped by rainfall and other climatic conditions (NMEP, 2021; USAIDs, 2019). Rainfall duration ranges from about three months in the Sahel savannah to nine months in the mangrove swamps and rainforest. These climatic patterns influence vegetation, creating diverse mosaics that differentiate flora, fauna, and mosquito vector species across zones (USAIDs, 2019).

For this study, five states are selected across the five ecological zones having sentinel sites where VectorLink Nigeria conducted longitudinal vector surveillance: Akwa-Ibom (South South, mangrove swamps/rainforest), Ebonyi (South East, rainforest), Kebbi (North West, Sahel Savannah), Oyo (South West, rainforest/Guinea-savannah) and Plateau (North Central, Guinea savannah) (PMI, 2022) (see Figure 1).

**Figure 1.**
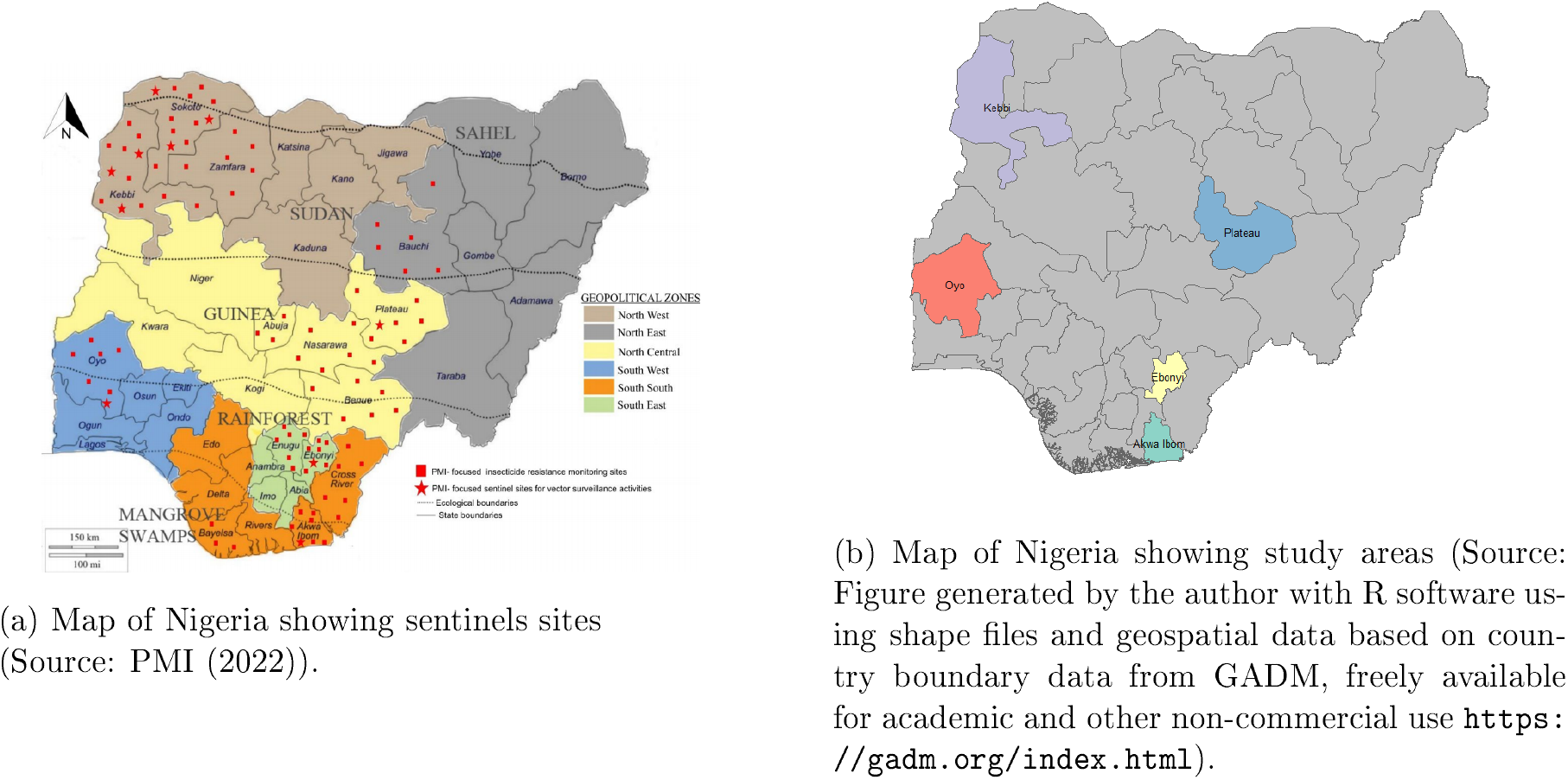
Maps of Nigeria showing sentinel sites (left pane) and selected study areas (right pane).

#### 2.1.1 Data

##### Monthly Malaria cases data

Nigeria Malaria Data Repository (NMDR) owned by Nigeria’s National Malaria Elimination Programme (NMEP) is a platform containing various malaria indicators from healthcare facilities across the country https://nmdrnigeria.ng/dhis-web-commons/security/login.action. On the platform are both routine (monthly) and non-routine (cross-sectional) data. For this research, monthly reported confirmed uncomplicated malaria cases from January 2015 to December, 2024 are obtained for each selected state through the NMDR platform. Figure 2 is the time series plot of confirmed uncomplicated malaria cases for each state being studied.

**Figure 2.**
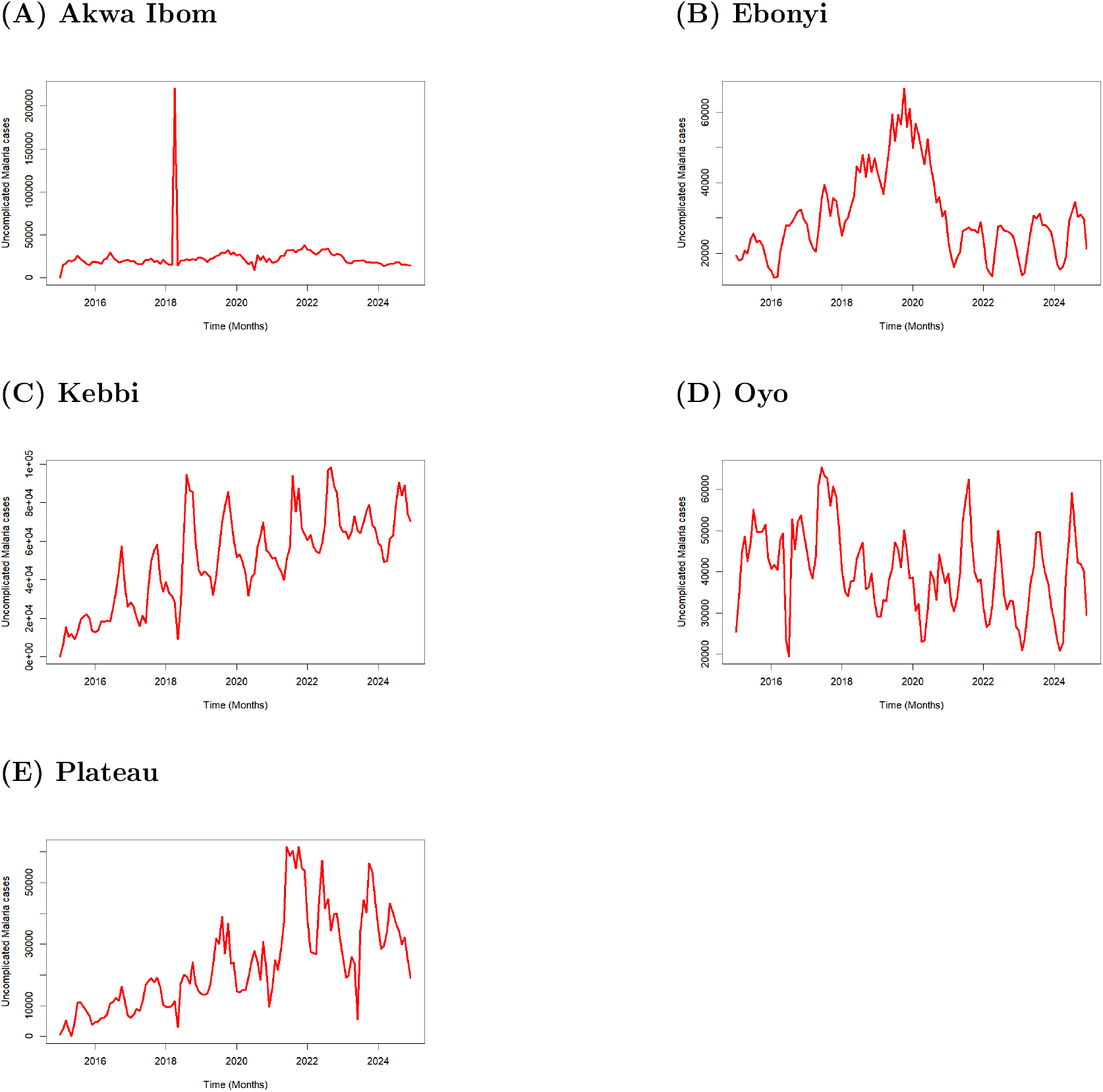
Time series plots of monthly confirmed uncomplicated malaria cases in Akwa Ibom, Ebonyi, Kebbi, Oyo and Plateau states from 2015 to 2024.

##### Demographic data

The National and State population projection based on 2006 Population and Housing Census (PHC) covering National, 36 States and FCT from 2007 to 2022 was carried out by the National Population Commission (NPC) (NPC, 2024). Total population across all states under study from 2015 to 2022 required for this study can be accessed https://nationalpopulation.gov.ng/publications.

### Malaria prevalence data

The Demographic and Health Surveys (DHS) Program funded by the United State Agency for International Development (USAID) implements demographic and health surveys and the Malaria Indicator Surveys (MIS) among other surveys. Available malaria prevalence data for 2015, 2018 and 2021 for all states under consideration are obtained from the DHS program platform through https://dhsprogram.com.

### Intervention data

Data on LLINs usage are obtained from the publicly available MIS reports. Readers are referred to NMEP (2016, 2022) which give detailed report of surveillance done to obtain data on various indicators of malaria including the use of LLINs across all the states in Nigeria.

Though malaria treatment is not the focus of this study, however, to cater for the reality on ground concerning malaria in the states under consideration, we made use of data on malaria treatment obtained from the NMDR platform at some point in this study.

### Entomological data

Entomological surveillance activities are being carried out by the President’s Malaria Initiative (PMI) from time to time in sentinels located at different states in Nigeria using human-baited Centers for Disease Control and Prevention light trap (CDC LT), (positioned indoors and outdoors) and Pyrethrum Spray Catch (PSC) methods (PMI, 2022). In all sentinels sites across all states under consideration, total number of anophelines caught (PSC and CDC LT collections), number of anophelines tested for sporozoites, number of anophelines tested positive for sporozoites, human biting rates of anophelines (Estimated from CDC LT collections) are extracted from reports of surveillance done from 2015 to 2022. For each of the survey, all variables of interest are summed over this survey period to obtained their total outcomes for the period under consideration. Tables 2 to 5 present the extracted variables. original reports can be accessed through https://www.pmi.gov.

### Data Preprocessing

Here, we address outliers to ensure data quality. For example, in the Akwa Ibom uncomplicated malaria case data shown in Figure 2, two points are identified as outliers because they are outside the typical data range. These points are removed, and their values are replaced with the mean of the remaining observations.

Additionally, we transform certain variables into a desired form compatible for use. For instance, using the number of anophelines caught indoors and outdoors across all sentinel sites in the states considered (Table 2), we estimate the population of female Anopheles mosquitoes present indoors or outdoors during the entomological surveillance periods conducted by PMI. This estimation is based on the formula:

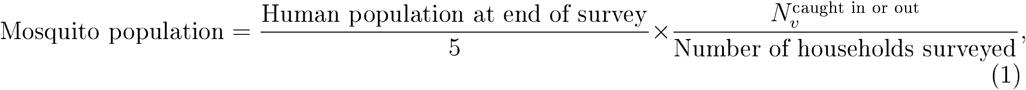

where 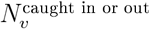 represents the number of anophelines caught either indoors or outdoors during the survey. We assumed an average of five persons per household, consistent with the 2015 and 2021 MIS (NMEP, 2016, 2021), so the first term gives the estimated number of households. The second term yields the average number of female *Anopheles* mosquitoes per household, and the product provides the estimated mosquito population indoors or outdoors, depending on the context of 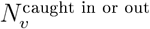.

### 2.2 Model formulation

#### 2.2.1 Description of model dynamics

Here, we formulate a deterministic compartmental model describing the transmission dynamics of malaria infection between humans and female *Anopheles* mosquitoes, incorporating realistic indoor and outdoor mosquito feeding settings and time-dependent LLIN effectiveness.

Based on infection status, human population is divided into susceptible *S*_*h*_, preinfectious *E*_*h*_, symptomatic infectious *I*_*h*_ and asymptomatic infectious *A*_*h*_ compartments. Total human population *N*_*h*_ is the aggregation of all the human classes, that is,

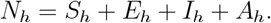

Female *Anopheles* mosquito population on the other hand is classified based on mosquito infection status and location of bite into indoor and outdoor biting susceptible female mosquitoes *S*_*vi*_ and *S*_*vo*_; indoor and outdoor biting pre-infectious female mosquitoes *E*_*vi*_ and *E*_*vo*_; indoor and outdoor biting infectious female mosquitoes *I*_*vi*_ and *I*_*vo*_. Total indoor biting female mosquitoes *N*_*vi*_, total outdoor biting female mosquitoes *N*_*vo*_ and total female mosquito population *N*_*v*_ are respectively given as:

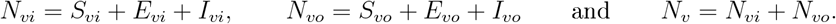

We assume that the proportion of human population that is effectively protected by LLIN usage at time *t, u*_*p*_(*t*), is a product of LLIN usage (proportion of the human population sleeping under the nets), efficacy of the nets for blood feeding inhibition and nets survival (measured as the proportion of the nets in functional state) at the given time. Suppose there are *n* LLIN mass distribution campaigns done so far in a state under consideration, let *u*_*i*_ be LLIN usage corresponding to the *i*^th^ campaign, for 1 ≤ *i* ≤ *n* and let *ω*_*p*_ be the blood feeding inhibition efficacy of LLINs at the time of acquisition. Given that *τ*_*i*_ is the time of *i*^th^ campaign (which is taken to be the time of LLIN acquisition), expression *t* − *τ*_*i*_ represents the time LLINs have been in use since the time of acquisition. From Obi et al. (2020), it is established that the proportion of the nets still in functional state decreases over time and that it reaches 50% at median survival time of the nets. Following a suggestion in Ngonghala et al. (2014), we use the exponential functional form 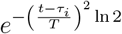 to represent the proportion of LLINs that is still in functional state at time *t*, with *T* being the median survival time of the nets. The rationales for choosing 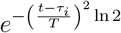 are first, that it is a smooth function bounded below and above by 0 and 1 respectively, hence it can be used to represent proportion; second, that the natural logarithm component can be used to moderate the proportion attained when *t* − *τ*_*i*_ = *T* which in this case is 50%, hence it captures the median survival of the nets; and third, that it is a decreasing function, hence it can be used to capture decay process that it is being engaged for. Therefore, effective protection *u*_*p*_(*t*) offered by LLIN usage across the duration of study [0, *T*_last_] is therefore given by

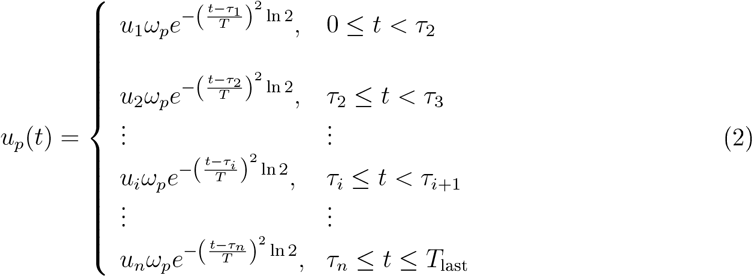

Observe from (2) that when *t* = *τ*_*i*_ then *u*_*p*_(*t*) = *u*_*i*_*ω*_*p*_: that is, at the time of acquisition, LLINs have full protective effectiveness. Also, when *t* − *τ*_*i*_ = *T*, then 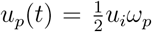: that is, when the period of time LLINs have been in use from the time of their acquisition is up to the median survival time, then the protective effectiveness of the net is half the initial level of protection at the time of acquisition. However, when *t* − *τ*_*i*_ *> T* and *t* → ∞, then *u*_*p*_(*t*) → 0: that is, as time goes on after the nets have been used beyond the median survival time, the protective effectiveness of the nets reduces until they are not be able to offer any protection again.

Now, a number of LLIN mass distribution campaigns have been done in Nigeria and the Nigeria Malaria Indicator Survey (MIS) with a primary objective of providing up-to-date estimates of basic demographic and health indicators associated with malaria is carried out every five years with one of the key information supplied by the survey being the estimate of mosquito nets usage across all states of the federation (NMEP, 2022). In order to align LLIN duration of use with LLIN mass distribution campaign year, we assume that LLIN usage reported by MIS in any year is due to the LLINs received at the earliest LLIN mass distribution campaign done before the survey year or just in the survey year. Since the monthly malaria cases data for this study are from year 2015 to 2024, we set the first day of January, 2015 as *t* = 0 to be our start time. It is possible that the earliest of MIS reported LLIN mass distribution campaigns comes before, after or just at time *t* = 0. That is, either *τ*_1_ *<* 0, *τ*_1_ > 0 or *τ*_1_ = 0 . When *τ*_1_ *<* 0, it means that the nets are acquired before the year 2015. For instance, if *τ*_1_ = −24, then the nets were acquired in the year 2013, which means that the nets have been in use two years before the start of study year. Hence, the nets are not expected to be in full capacity of effectiveness at 2015 due to loss of physical integrity. For the case of *τ*_1_ = 0, the nets are acquired at the start time, hence they will be able to offer full protection at the start. Lastly, for the situation where *τ*_1_ > 0, the nets were distributed after the start time, so it is assumed no one is under LLIN protection at the start of the study, so *u*_*p*_(*t*) = 0 till the time when campaign is carried out. For instance, if *τ*_1_ = 12 (no campaign till a year after the start, that is year 2016), then *u*_*p*_(*t*) = 0 until year 2016. Given that *u*_1_ = 0.38, *u*_2_ = 0.48 and *ω*_*p*_ = 0.98, Figure 3 gives a graphical illustration of cases with *τ*_1_ = −24 (year 2013), *τ*_1_ = 0 (year 2015) and *τ*_1_ = 12 (year 2016) with corresponding *τ*_2_ being *τ*_2_ = 36 (year 2018), *τ*_2_ = 48 (year 2019) and *τ*_2_ = 60 (year 2020) respectively.

**Figure 3.**
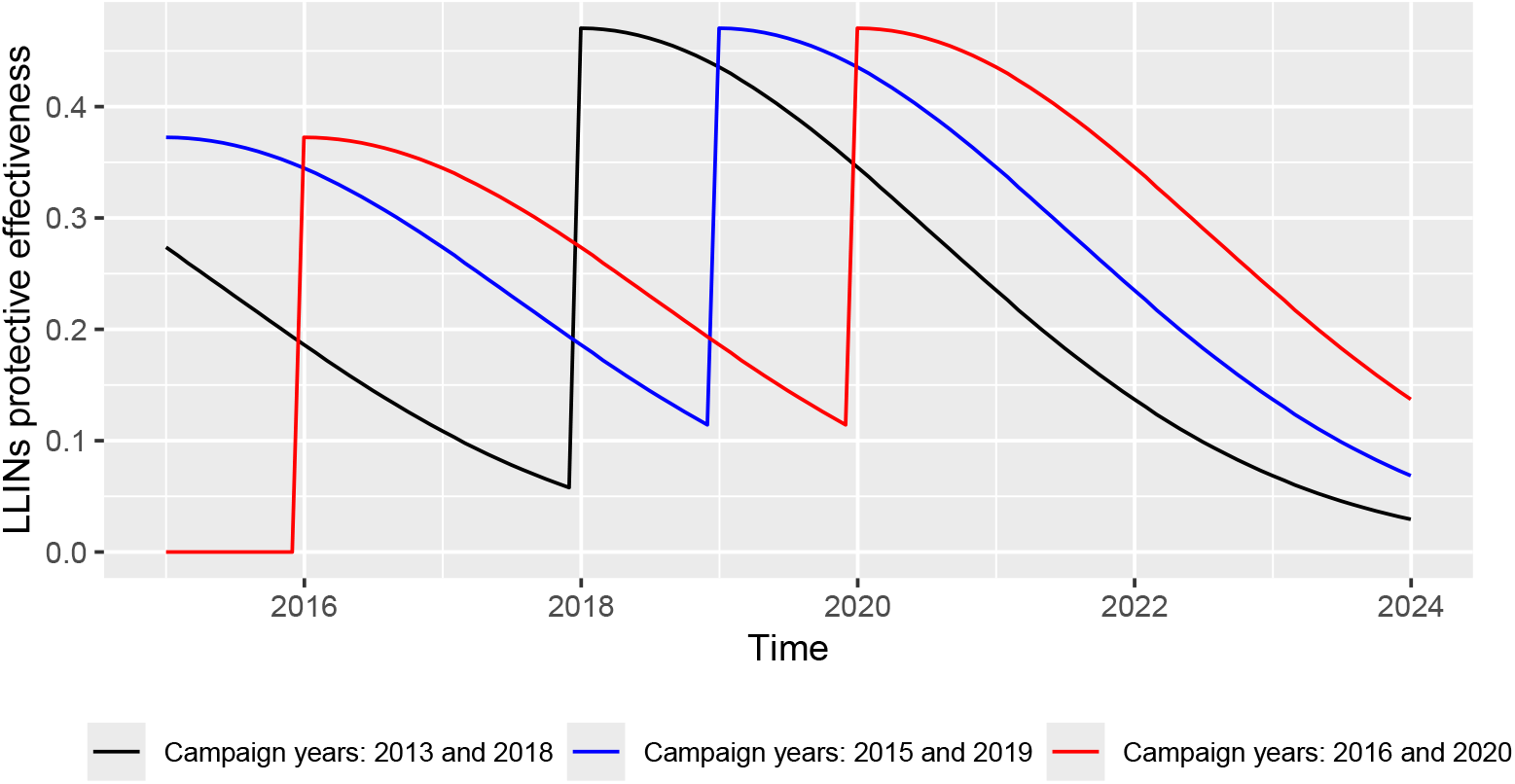
Plot illustrating LLIN protective effectiveness at time *t, u*_*p*_(*t*) in (2) based on different times of LLIN acquisition (campaign years).

Since it is as mosquitoes attempt biting a person protected by LLINs that they get killed, as biting attempt increases, mosquito mortality due to LLIN usage also increases. Given *u*_*k*_(*t*) as the killing effectiveness of LLINs (proportion of mosquitoes that die due to contact with LLINs), *ε*_*vi*_ the mean indoor blood feeding rates of mosquitoes, *a* the proportion of mosquito indoor blood meal taken in-bed, mosquito mortality rate due to LLIN usage *µ*_*k*_(*t*) at time *t* is assumed to be a product of indoor in-bed feeding rate of mosquitoes and the killing effectiveness of LLINs, that is

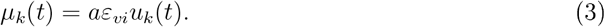

Similar to the case of effective protection offered by LLINs to its users, the mosquitoes killing effect of LLINs, *u*_*k*_(*t*), that can be achieved in a population is also assumed to be the product of LLIN usage, insecticidal efficacy of LLINs and nets survival. Though, it is shown in Obi et al. (2020) that the LLIN distributed during the 2015/16 mass distribution campaign (DAWA plus 2.0) have optimal insecticidal performance based on WHO criteria after three years of use, we reason that nets survival will still mediate in the killing capacity of the nets, since it is only the surviving nets that can kill mosquitoes given that the insecticide used is only embedded in the nets and thus can not act independently of the nets. Suppose *u*_*i*_, *τ*_*i*_, *T* are as defined before and *ω*_*k*_ is the insecticidal efficacy of newly acquired LLINs, then the killing effectiveness of LLINs *u*_*k*_(*t*) at time *t* is given as:

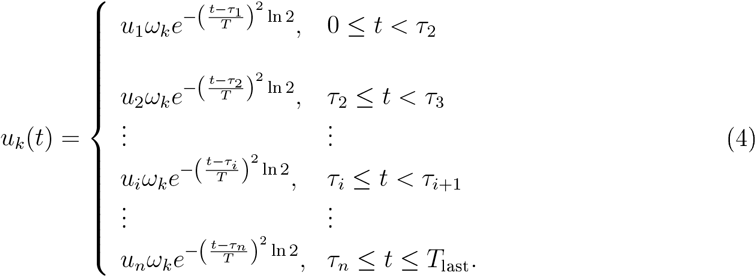

The force of infection in human is derived based on the assumptions that people use LLINs indoors while in-bed only. Thus given that *ε*_*vo*_ and *ε*_*vi*_ are respectively the mean outdoor and indoor blood feeding rates of mosquitoes, *a* is the proportion of mosquito indoor blood meal taken in-bed while (1 − *a*) is the proportion of indoor blood meal taken out of bed and *β*_*h*_ is the probability of successful infection transmission in human after mosquito bite assumed to be the same indoors and outdoors, then force of infection in human population is given as:

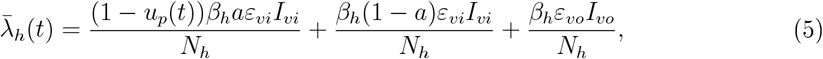

where the products *β*_*h*_*ε*_*vi*_ and *β*_*h*_*ε*_*vi*_ give the average transmission rates in humans due to indoor and outdoor blood feeding rates of mosquitoes respectively, and the three terms in (5) represent forces of infection due to indoor in-bed, indoor out of bed and outdoor blood feeding rates of mosquitoes respectively. This implies that total malaria infection that occur in a population is the sum of malaria infections that occur indoors in-bed, indoors out of bed and outdoors. To account for the periodic fluctuations exhibited by the observed malaria cases data, Fourier series is incorporated into the force of infection 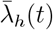 so that

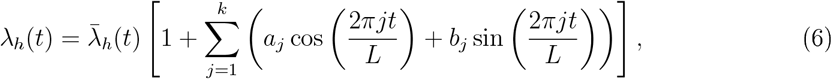

where *a*_*j*_ and *b*_*j*_ are coefficients of the *j*^th^ harmonic of the Fourier series, *L* is the period and *k* is the suitable number of harmonics.

Also,we assume that upon successful blood meal from either symptomatic or asymptomatic infectious humans, indoor and outdoor host seeking mosquitoes become infected indoors and outdoors with forces of infection 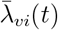 and 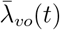 respectively, such that

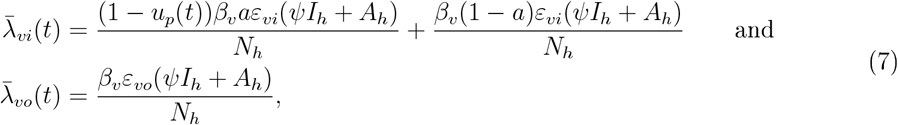

where the two terms of 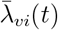 represent forces of infection in mosquitoes due to indoor in-bed and indoor out of bed biting respectively, *β*_*v*_ is the probability of successful transmission in mosquitoes after a successful blood meal either indoor or outdoor, and *ψ* is a modification parameter such that 0 *< ψ <* 1, arising as a result of reduced level of gametocytes in symptomatic infectious individuals (Lindblade et al., 2013). Similar to what we did in the human population, the forces of infection 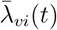 and 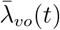 are multiplied by a Fourier series, so that,

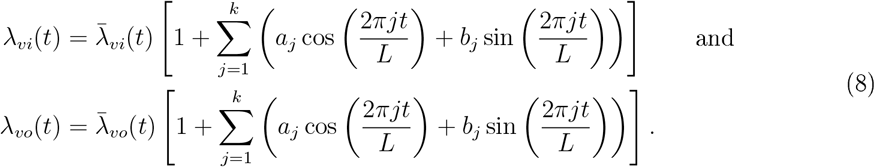

The susceptible human class *S*_*h*_ is increased by human recruitment which is assumed to be at constant rate Λ_*h*_ and addition of symptomatic infectious individuals who recover from an episode of previous malaria infection due to treatment at rate *γ*. Here, it is assumed that symptomatic individuals recover to join the susceptible human class since in an endemic setting, an individual does not acquire infection blocking immunity after recovery, but can rather get infected almost immediately if exposed to infectious bite from female *Anopheles* mosquito (Mandala et al., 2021). Also, the class is reduced through natural death at rate *µ*_*h*_ and force of infection *λ*_*h*_(*t*) due to infectious bite of female *Anopheles* mosquitoes either indoors or outdoors. Hence, the differential equation representing the dynamics of the susceptible human population is therefore given by:

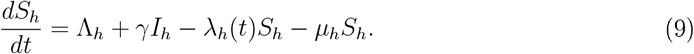

Now, the pre-infectious human class *E*_*h*_ which contains individuals who are infected but are yet to be infectious is increased owing to susceptible individuals who become infected due to the force of infection *λ*_*h*_(*t*). Also, the class decreases through individuals in the class who either die naturally at rate *µ*_*h*_ or move to the infectious human class at per capital rate *σ*_*h*_. Hence, the differential equation representing the dynamics of the pre-infectious human class is given by:

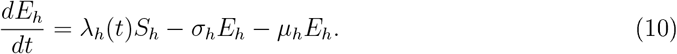

Next, the symptomatic infectious human class *I*_*h*_ increases when individuals from the pre-infectious human class become infectious with onset of symptoms at rate *p*_1_*σ*_*h*_, where *p*_1_ is the probability that symptoms will appear at the end of latent period 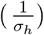. We assume that with more infectious bite, an asymptomatic infectious human can become symptomatic infectious at a rate *p*_2_*λ*_*h*_(*t*), where *p*_2_ is the probability of an asymptomatic individual becoming symptomatic leading to an increase in the class. However, the class decreases as a result of natural death at rate *µ*_*h*_, recovery from the disease at per capital recovery rate *γ* due to treatment and additional death due to the disease at per capital disease induced death rate *δ*_*h*_. All these then culminate into the differential equation:

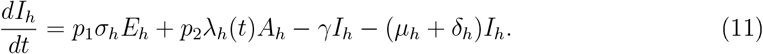

Individuals are added to the asymptomatic infectious class *A*_*h*_ from the pre-infectious human class at rate (1 − *p*_1_)*σ*_*h*_ with probability 1 − *p*_1_ of being asymptomatic at the end of latent period. Also, the class reduces due natural death at rate *µ*_*h*_ and more infectious bite leading to movement to symptomatic class at rate *p*_2_*λ*_*h*_(*t*). Asymptomatic individual are assumed not to die as a result of the disease. Hence, the dynamics of the asymptomatic infectious human class is given by the differential equation

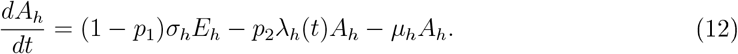

Just as in the case of human population, susceptible female *Anopheles* mosquitoes are assumed to be recruited constant rate Λ_*v*_ with an assumption that proportion *q* of the recruited susceptible female *Anopheles* mosquitoes find their way indoors for host seeking while proportion 1 − *q* are outdoor host seeking susceptible mosquitoes.

Moreover, indoor susceptible mosquitoes move outdoors at rate *α*_*io*_ and outdoor susceptible mosquitoes move indoors at rate *α*_*oi*_. Indoor and outdoor susceptible mosquitoes die naturally at rate *µ*_*v*_ while indoor mosquitoes that attempt biting a person protected by LLINs in-bed at time *t* suffer an additional death due to the killing effectiveness of LLINs’ insecticidal property at rate *µ*_*k*_(*t*).

The differential equations representing the dynamics of indoor and outdoor susceptible female *Anopheles* mosquito populations are therefore respectively by:

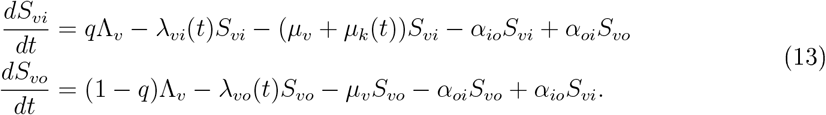

Indoor and outdoor mosquitoes move from susceptible class to pre-infectious class due to forces of infection *λ*_*vi*_(*t*) and *λ*_*vo*_(*t*) respectively. Indoor pre-infectious mosquitoes move outdoors at rate *α*_*io*_ and outdoor pre-infectious mosquitoes move indoors at rate *α*_*oi*_. Indoor and outdoor pre-infectious mosquitoes die naturally at rate *µ*_*v*_ while indoor pre-infectious mosquitoes that attempt biting a person protected by LLINs in-bed at time *t* suffer an additional death due to the killing effectiveness of LLINs’ insecticidal property at rate *µ*_*k*_(*t*). Indoor and outdoor pre-infectious mosquitoes become infectious at per capital extrinsic incubation rate *σ*_*v*_. Then, the differential equation governing the dynamics of indoor and outdoor pre-infectious mosquitoes are respectively given by:

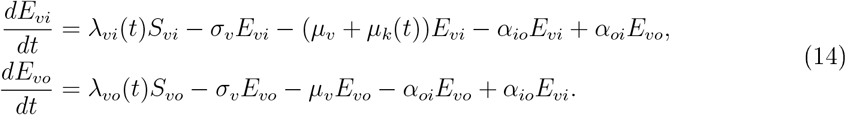

Finally, indoor and outdoor pre-infectious mosquitoes respectively move to indoor and outdoor infectious classes at rate *σ*_*v*_. Indoor infectious mosquitoes move outdoors at rate *α*_*io*_ and outdoor infectious mosquitoes move indoors at rate *α*_*oi*_. Indoor and outdoor infectious mosquitoes die naturally at rate *µ*_*v*_ while indoor infectious mosquitoes that attempt biting a person protected by LLINs in-bed at time *t* suffer an additional death due to the killing effectiveness of LLINs’ insecticidal property at rate *µ*_*k*_(*t*). It is assumed that infectious mosquitoes do not suffer disease induced death. The following differential equations consequently represent the dynamics of indoor and outdoor infectious mosquito populations:

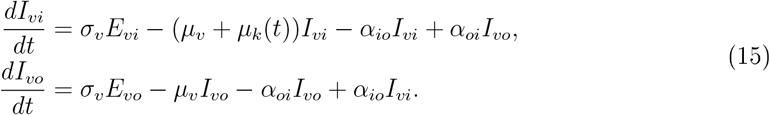

Schematic diagram representing the model described is given in Figure 4.

**Figure 4.**
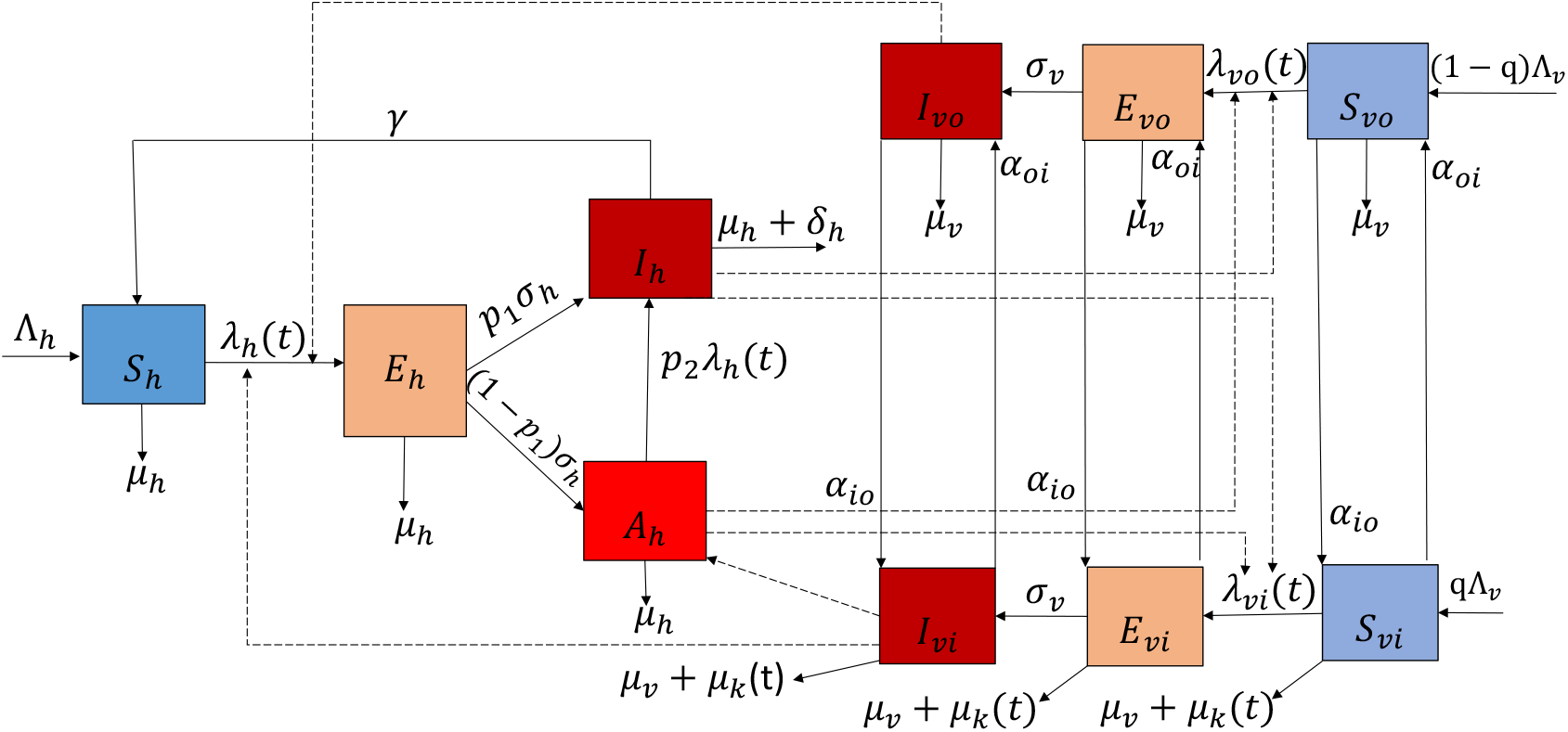
Schematic diagram for model (16).

Combining equations (9) to (15), we obtain the model representing the transmission dynamics of malaria infection between human and female Anopheles mosquito populations as the system of ordinary differential equations (ODEs) :

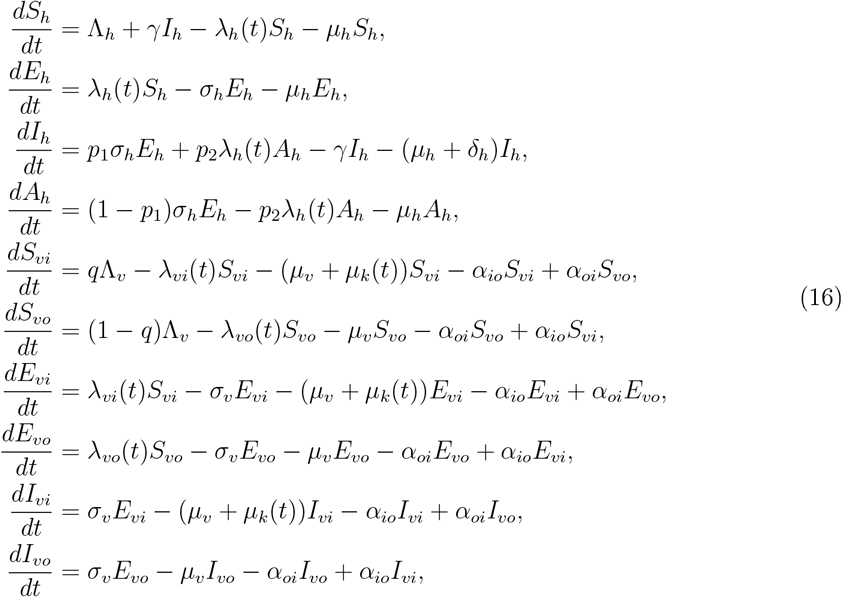

with initial conditions *S*_*h*_(0) > 0, *E*_*h*_(0) ≥ 0, *I*_*h*_(0) > 0, *A*_*h*_(0) > 0, *S*_*vo*_(0) > 0, *S*_*vi*_(0) > 0, *E*_*vi*_(0) ≥ 0, *E*_*vo*_(0) ≥ 0, *I*_*vi*_(0) > 0, *I*_*vo*_(0) > 0.

Table 6 and Table 7 are the descriptions of model variables and parameters respectively.

### 2.3 Quantitative Analysis

#### 2.3.1 Model calibration

Model calibration involves model fitting to data and parameter estimation. Calibrating model (16) will enable us to customize the model for each of the states under consideration in this study. To do this, initial values for the state variables are required together with values for those parameters that would not be estimated.

##### Initial conditions

The initial conditions for all the state variables are hereby stated: Based on the reported uncomplicated malaria cases, *I*_*h*_(0) is taking to be the number of malaria cases reported by January 2015 for each of the states, therefore, *I*_*h*_(0) = 22 197 (Akwa Ibom), 19 336 (Ebonyi), 177 (Kebbi), 25 517 (Oyo), and 577 (Plateau). It is believed that in endemic areas, for every symptomatic malaria case, there are 4 to 5 asymptomatic carriers (Abebaw et al., 2022), hence we assume that *A*_*h*_(0) = 5*I*_*h*_(0), so that *A*_*h*_(0) = 110 985 (Akwa Ibom), 96 680 (Ebonyi), 885 (Kebbi), 127 585 (Oyo) and 2 885 (Plateau). We assume *E*_*h*_(0) = 0 for all states.

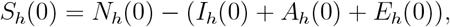

and using *N*_*h*_(0) as 2015 human population for each state from Table 1, we have *S*_*h*_(0) = 4 373 862 (Akwa Ibom), 2 599 659 (Ebonyi), 4 376 708 (Kebbi), 6 745 694 (Oyo) and 4 001 944 (Plateau). *I*_*vi*_(0) and *I*_*vo*_(0) are obtained using

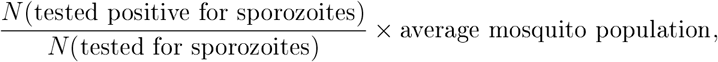

where *N* (tested for sporozoites) is the sum of all female *Anopheles* mosquitoes tested for sporozoites across all surveys done in each state and *N* (tested positive for sporozoites) is the corresponding sum of those that tested positive, so that the quotient 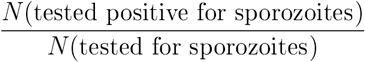 gives the proportion of the tested mosquitoes that are positive for sporozoites and average mosquito population per survey is obtained by dividing the sum of mosquito population estimated throughout all surveys using (1) by the number of surveys. This process is carried out for both indoor and outdoor context to obtain *I*_*vi*_(0) and *I*_*vo*_(0) respectively and this yielded *I*_*vi*_(0) = 965 657 (Akwa Ibom), 938 520 (Ebonyi), 3 436 798 (Kebbi), 2 656 329 (Oyo) and 1 173 821; and *I*_*vo*_(0) = 1 332 838 (Akwa Ibom), 585 628 (Ebonyi), 4 171 795 (Kebbi), 4 291 638 (Oyo) and 1 912 092 (Plateau). We assume *E*_*vi*_(0) = *E*_*vo*_(0) = 0 for all states. *S*_*vi*_(0) = *N*_*vi*_(0) − (*I*_*vi*_(0) + *E*_*vi*_(0)) and *S*_*vo*_(0) = *N*_*vo*_(0) − (*I*_*vo*_(0) + *E*_*vo*_(0)), where *N*_*vi*_(0) and *N*_*vo*_(0) are taken as the earlier discussed average mosquito population indoor and outdoor respectively, so that *S*_*vi*_(0) = 41 705 934 (Akwa Ibom), 29 273 822 (Ebonyi), 721 727 533 (Kebbi), 92 848 907 (Oyo) and 102 826 680 (Plateau); and *S*_*vi*_(0) = 75 471 951 (Akwa Ibom), 17 158 911 (Ebonyi), 826 750 402 (Kebbi), 111 808 475 (Oyo) and 90 015 398 (Plateau).

**Table 1:**
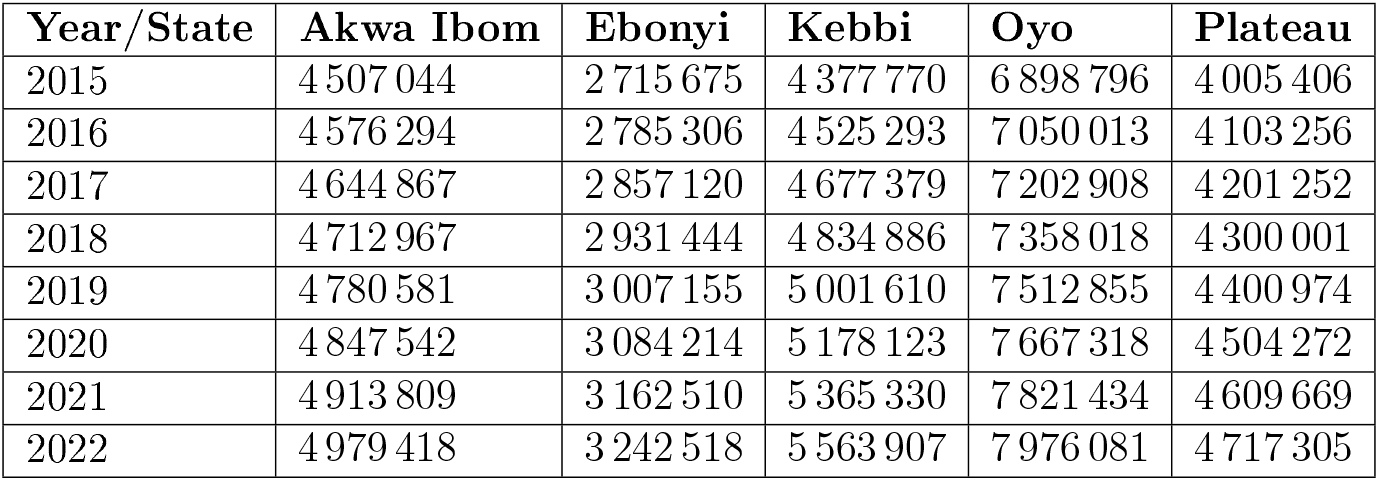
Population projection from 2015 to 2022 for states under study (extracted from NPC (2024))

**Table 2:** Total number of anophelines caught in all sentinel sites(PSC and CDC LT collections)(extracted from PMI (2015, 2016, 2017, 2018, 2019, 2020, 2021, 2022))

| State/Date | Nov<br>14-Dec<br>15 | Feb-Dec<br>16 | Jan-Dec<br>17 | Apr-Sep<br>18 | Nov<br>18-Sep<br>19 | Oct<br>19-Sep<br>20 | Oct<br>20-Sep<br>21 | Oct<br>21-Sep<br>22 |
| --- | --- | --- | --- | --- | --- | --- | --- | --- |
| Akwa-Ibom In |  | 1 656 | 1 593 | 952 | 2 382 | 1 570 | 1 334 | 1 757 |
| Akwa-Ibom Out |  | 663 | 528 | 97 | 115 | 364 | 228 | 276 |
| Ebonyi In |  | 3 141 | 1 761 | 1 594 | 1 556 | 1 455 | 1 774 | 1 492 |
| Ebonyi Out |  | 115 | 151 | 131 | 348 | 41 | 35 | 20 |
| Kebbi In |  |  |  |  |  |  | 12 213 | 35 143 |
| Kebbi Out |  |  |  |  |  |  | 2 259 | 3 790 |
| Oyo In |  | 894 | 759 | 557 | 4 212 | 4 380 | 3 719 | 1 273 |
| Oyo Out |  | 74 | 41 | 13 | 593 | 689 | 540 | 173 |
| Plateau In | 3 624 |  |  | 4 515 | 5 587 | 4 486 | 3 512 | 3 509 |
| Plateau Out | 501 |  |  | 220 | 442 | 577 | 182 | 146 |

**Table 3:**
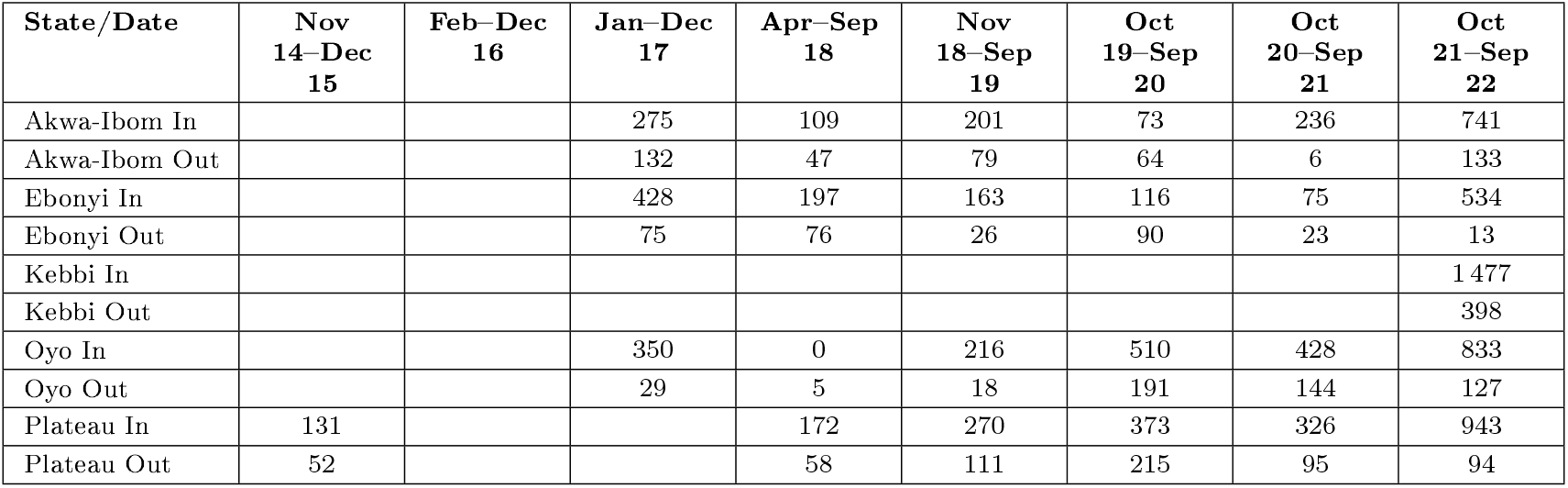
Number of Anophelines tested for sporozoites in all Sentinel sites (extracted from PMI (2015, 2016, 2017, 2018, 2019, 2020, 2021, 2022))

**Table 4:** Number of Anophelines tested positive for sporozoites in all Sentinel sites (extracted from PMI (2015, 2016, 2017, 2018, 2019, 2020, 2021, 2022))

| State/Date | Nov<br>14-Dec<br>15 | Feb-Dec<br>16 | Jan-Dec<br>17 | Apr-Sep<br>18 | Nov<br>18-Sep<br>19 | Oct<br>19-Sep<br>20 | Oct<br>20-Sep<br>21 | Oct<br>21-Sep<br>22 |
| --- | --- | --- | --- | --- | --- | --- | --- | --- |
| Akwa-Ibom In |  |  | 24 | 6 | 2 | 1 | 1 | 3 |
| Akwa-Ibom Out |  |  | 7 | 1 | 0 | 0 | 0 | 0 |
| Ebonyi In |  |  | 31 | 8 | 2 | 1 | 1 | 4 |
| Ebonyi Out |  |  | 7 | 2 | 1 | 0 | 0 | 0 |
| Kebbi In |  |  |  |  |  |  |  | 7 |
| Kebbi Out |  |  |  |  |  |  |  | 2 |
| Oyo In |  |  | 23 | 0 | 15 | 5 | 20 | 2 |
| Oyo Out |  |  | 7 | 0 | 0 | 0 | 9 | 3 |
| Plateau In | 8 |  |  | 5 | 2 | 0 | 7 | 3 |
| Plateau Out | 1 |  |  | 10 | 1 | 0 | 1 | 0 |

##### Sourced parameters

The procedural steps taken to obtain the parameter values presented in Table 8 are given here. Some of the model parameter values will be obtained either directly or derived based on information from literature. Human recruitment rate Λ_*h*_, is obtained for each state using

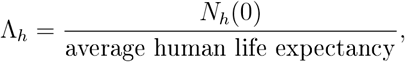

where *N*_*h*_(0) is as ealier given for each state and average human life expectancy for each state is calculated by dividing the sum of average life expectancy for males and females in 2015 from NPC (2024) by 2, then the result is divided by 12 to have the outcome in monthly time unit. Using the described approach, we have Λ_*h*_ = 6 712.9 (Akwa Ibom), 4 129.676 (Ebonyi), 6 456.89 (Kebbi), 10 275.24 (Oyo) and 6 702.487 (Plateau) all per month. Natural mortality rate per month is obtained by taking the reciprocal of average human life expectancy in month, hence *µ*_*h*_ = 1.489 × 10^*−*3^ per month (Akwa Ibom), 1.521 × 10^*−*3^ per month (Ebonyi), 1.475 × 10^*−*3^ per month (Kebbi), 1.489 × 10^*−*3^ per month (Oyo) and 1.673 × 10^*−*3^ per month (Plateau). Progression rate from pre-infectious class to infectious classes in human, *σ*_*h*_ is derived as the reciprocal of latent period of *P. falciparum*. Based on the work of Chitnis et al. (2008), we use 10 days as the latent period, so that *σ*_*h*_ = 3 per month for all states. Recovery rate due to treatment is obtained using

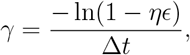

where *η* is the treatment coverage (proportion of confirmed uncomplicated malaria cases treated with ACT), *ϵ* is the efficacy of ACT, and Δ*t* is the time it takes ACT to clear malaria infection. The product *ηϵ* gives the proportion of individuals whose infection get cleared due to treatment with ACT (probability of infection clearance). The derivation of *γ* is a consequence of the assumption that the malaria infection clearance follows an exponential distribution and the probability of infection clearance, *ηϵ* = 1−exp(−*γ*Δ*t*). Using the columns representing number of uncomplicated malaria cases and number of uncomplicated malaria cases treated with ACT from the NMDR platform, we obtain *η* for all states using

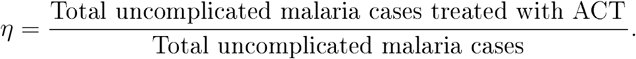

Thus, *η* = 0.983 (Akwa Ibom), 0.995 (Ebonyi), 0.992 (Kebbi), 0.993 (Oyo) and 0.966 (Plateau). In Nigeria, reports of drug therapeutic efficacy tests/trials (DTETs) have shown adequate clinical and parasitological response (ACPR) (defined as the absence of parasitemia on day 28) of about 98% for first-line ACT treatments (Ojurongbe et al., 2013; NMEP et al., 2018; NMEP, 2021), hence we take *ϵ* = 0.98 for all states. Using the fact that the primary efficacy end-point of DTETs was a day 28 polymerase chain reaction (PCR)-corrected parasitological cure (Ojurongbe et al., 2013), we set Δ*t* = 28 days = 0.933 month for all states. Therefore, *γ* is explicitly determined for all states as *γ* = 3.547 per month (Akwa Ibom), 3.957 per month (Ebonyi), 3.839 per month (Kebbi), 3.875 per month (Oyo) and 3.148 per month (Plateau). As earlier stated that in endemic regions, for every symptomatic person there are 4 to 5 asymptomatic individuals (Abebaw et al., 2022), assuming ratio 1 : 5, we have that the proportion *p*_1_ of individuals progressing from pre-infectious class to symptomatic class is 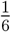 while 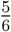 progress to asymptomatic class. In Lindblade et al. (2013), it is stated that, asymptomatic individuals transmit 61% more the gametocytes level than symptomatic individuals. This implies that whatever level of gametocytes transmitted by a typical symptomatic individual, an asymptomatic individual transmits 161% of that level of gametocytes, hence, we can say that whatever level of gametocytes an asymptomatic individual transmits, a symptomatic individual transmits 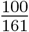 of that gametocytes level. Therefore, we set the modification parameter 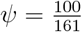. Mosquito recruitment rate Λ_*v*_ is obtained in a similar way to human recruitment rate as

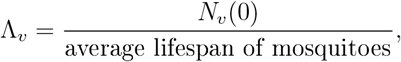

where *N*_*v*_(0) = *N*_*vi*_(0) + *N*_*vo*_(0) and average lifespan of mosquitoes is taking to be 12 days ( 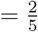 month) using the average of the range reported in CDC (2024). Then, Λ_*v*_ = 2.99 × 10^8^ per month (Akwa Ibom), 1.199 × 10^8^ per month (Ebonyi), 3.89 × 10^9^ per month (Kebbi), 5.29 × 10^8^ per month (Oyo) and 4.898 × 10^8^ per month (Plateau). Per capital natural death rate of mosquitoes *µ*_*v*_ = 1*/*average lifespan of mosquitoes = 5*/*2 per month. The rate of progression from pre-infectious class of female *Anopheles* mosquitoes to infectious class *σ*_*v*_ is obtained as the reciprocal of latent period for sporozoites in mosquitoes which is 14 days (14*/*30 month) (CDC, 2024), so that *σ*_*v*_ = 15*/*17 per month. Proportion of susceptible mosquitoes that seek blood meal indoors, *q* is obtained using the fraction

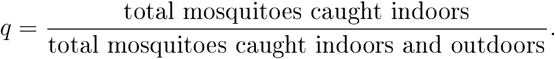

Hence, *q* = 0.357 (Akwa Ibom), 0.63 (Ebonyi), 0.466 (Kebbi), 0.451 (Oyo) and 0.531 (Plateau). As a proxy, proportion of mosquitoes indoor blood meal taken in-bed *a* is determined by using the fraction of PMI survey time (6:00 pm to 6:00 am) that people spend indoor in-bed. We assume that on average, people go to bed by 10:00 pm hence *a* = 2*/*3. Suppose *ε*_*hi*_ is the average human indoor biting rate and *ε*_*ho*_ is the average human outdoor biting rate, also, given that average mosquito outdoor and indoor blood feeding rates are *ε*_*vo*_ and *ε*_*vi*_ respectively, we can express *ε*_*hi*_ in terms of *ε*_*vi*_ and *ε*_*ho*_ in terms of *ε*_*vo*_: assuming the total number of blood meals taken per time by all mosquitoes in a place indoors (or outdoors) balance with the total number of mosquito bites received per time by individuals indoors (or outdoors) in that place i.e.

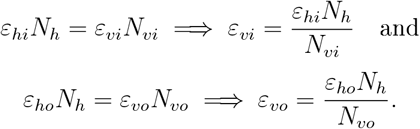

Table 5 is the report of human indoor and outdoor biting rates from PMI given in numbers of bite per human per day. We found the average human biting rate across all the survey period and convert it to monthly mosquito biting rates using the expressions for *ε*_*vi*_ and *ε*_*vo*_ multiplied by 30, where *N*_*vi*_, *N*_*vo*_ and *N*_*h*_ are taken as *N*_*vi*_(0), *N*_*vo*_(0) and *N*_*h*_(0) respectively. Hence, *ε*_*vi*_ = 12.04740 per month (Akwa Ibom), 10.3539 per month (Ebonyi), 2.1452 per month (Kebbi), 9.3738 per month (Oyo) and 21.4178 per month (Plateau); *ε*_*vo*_ = 3.2392 per month (Akwa Ibom), 3.6462 per month (Ebonyi), 0.6851 per month (Kebbi), 2.5264 per month (Oyo) and 2.3975 per month (Plateau). Using 2015 mortality rate of malaria from MAP (2022), we have *δ*_*h*_ = 7.454 × 10^*−*5^ per month (Akwa Ibom), 8.264 × 10^*−*5^ per month (Ebonyi), 1.24 × 10^*−*5^ per month (Kebbi), 9.285 × 10^*−*5^ per month (Oyo) and 7.313 × 10^*−*5^ per month (Plateau). With January, 2015 used a referenced time (*t* = 0), time of acquisition of LLINs, *τ*_*i*_ (1 ≤ *i* ≤ *n*, where n is the number of LLIN campaigns done) prior to, at and after time *t* = 0 takes negative value, zero and positive value respectively. Each of the states being considered has carried out LLIN campaign two time between 2009 and 2024 (NMEP and NPC and ICF, 2022), hence *n* = 2 for all states. Therefore *τ*_1_ = −12 and *τ*_2_ = 36 (Akwa Ibom), *τ*_1_ = 0 and *τ*_2_ = 48 (Ebonyi), *τ*_1_ = 0 and *τ*_2_ = 36 (Kebbi), *τ*_1_ = 12 and *τ*_2_ = 72 (Oyo), *τ*_1_ = 0 and *τ*_2_ = 60 (Plateau). Following the WHO acceptance criteria for LLINs (mortality of mosquitoes ≤ 80% or blood-feeding inhibition ≤ 90%), we set the protective effectiveness of LLINs *ω*_*p*_ = 0.90 and the killing effectiveness of LLINs *ω*_*p*_ = 0.8. LLIN usage is assumed to be as a result of LLINs acquired during LLIN mass campaign, hence we take LLIN usage reported in any MIS as one corresponding to the immediate past campaign except for Oyo state where *τ*_1_ = 12 indicating that LLIN campaign was done one year (2016) after the base year of this study, however, MIS was conducted in 2015, so we assume that LLIN usage reported in MIS 2015 is the same as LLIN usage corresponding to 2016 LLIN mass campaign. Therefore, *u*_1_ = 0.368 and *u*_2_ = 0.178 (Akwa Ibom), *u*_1_ = 0.494 and *u*_2_ = 0.481 (Ebonyi), *u*_1_ = 0.376 and *u*_2_ = 0.382 (Kebbi), *u*_1_ = 0.314 and *u*_2_ = 0.312 (Oyo), *u*_1_ = 0.384 and *u*_2_ = 0.267 (Plateau). The median survival time for LLINs was determined using the 2019 report of net durability monitoring study conducted in Nigeria, where Ebonyi, Oyo and Zamfara states were the three selected states (Obi et al., 2020). Assuming that the conditions responsible for the survivability of bed-nets are the same in somewhat similar ecological zones, we use the result from Ebonyi (rainforest ecological zone) for Akwa Ibom state (mangrove/rainforest ecological zone) and the result from Zamfara for Kebbi and Plateau states since they are all in the savannahs. Therefore, *T* = 39.6 months (Akwa Ibom and Ebonyi), 38.4 months (Oyo) and 63.3 months (Kebbi and Plateau). Finally, we set *T*_*last*_ = 120 being the number of months for the duration of study.

**Table 5:** Biting rates of *Anopseles* mosquitoes (estimated from CDC LT collections), extracted from PMI (2015, 2016, 2017, 2018, 2019, 2020, 2021, 2022)

| State/Date | Nov<br>14–Dec<br>15 | Feb–Dec<br>16 | Jan–Dec<br>17 | Apr–Sep<br>18 | Nov<br>18–Sep<br>19 | Oct<br>19–Sep<br>20 | Oct<br>20–Sep<br>21 | Oct<br>21–Sep<br>22 |
| --- | --- | --- | --- | --- | --- | --- | --- | --- |
| Akwa Ibom (In) |  |  |  | 46.91 | 59.59 | 82.75 | 7.18 | 5.08 |
| Akwa Ibom (Out) |  |  | 70.9 | 10.63 | 18.49 | 18.16 | 0.92 | 0.50 |
| Ebonyi (In) |  |  |  | 148.32 | 79.58 |  | 49.18 | 51.57 |
| Ebonyi (Out) |  |  | 24.2 | 52.93 | 7.17 |  | 2.57 | 1.00 |
| Kebbi (In) |  |  |  |  |  |  |  | 142.14 |
| Kebbi (Out) |  |  |  |  |  |  |  | 51.97 |
| Oyo (In) |  |  |  | 0.00 | 53.59 | 45.49 | 99.13 | 31.05 |
| Oyo (Out) |  |  | 7.00 | 0.00 | 28.75 | 8.75 | 33.15 | 14.47 |
| Plateau (In) |  |  |  | 78.19 | 181.35 | 211.00 | 178.03 | 158.05 |
| Plateau (Out) |  |  |  | 18.29 | 22.66 | 31.76 | 13.85 | 10.65 |

**Table 6:**
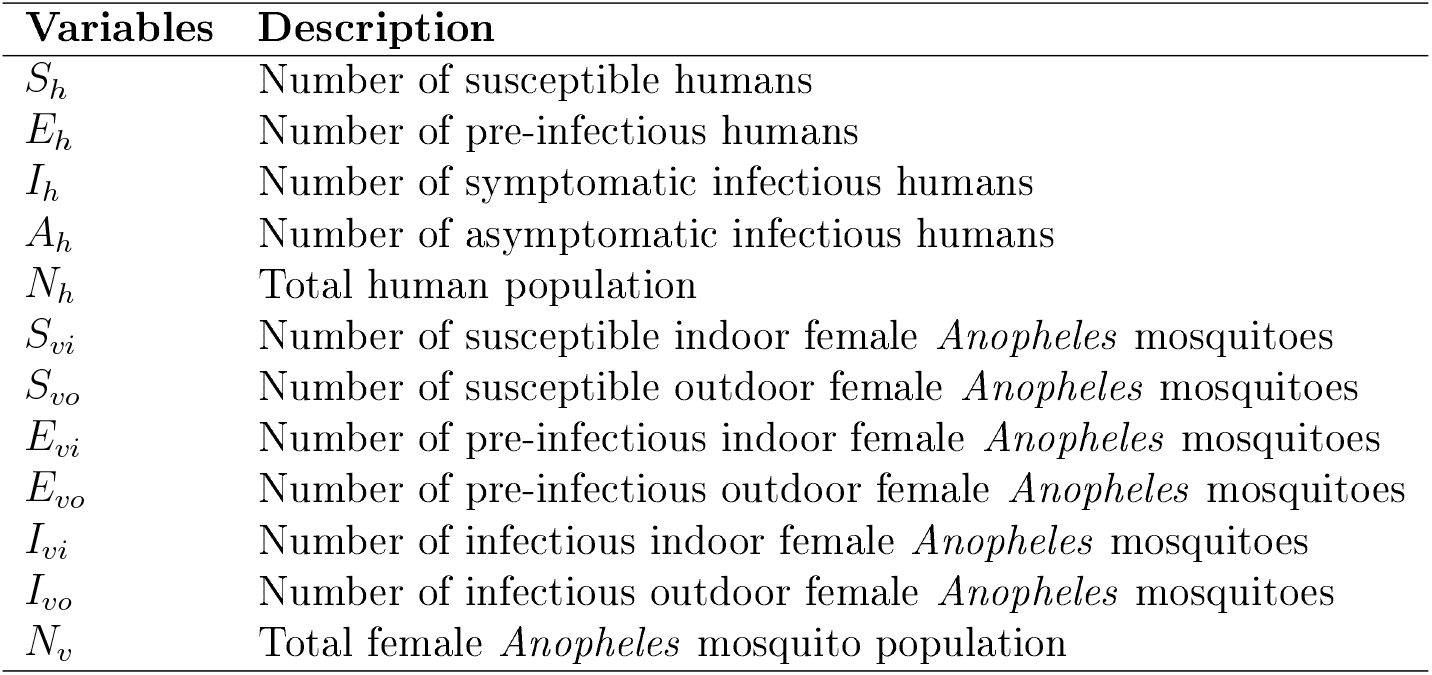
Description of model variables.

**Table 7:** Description of model parameters.

| Parameters | Description | Unit |
| --- | --- | --- |
| $\Lambda_h$ | Human recruitment rate | people × month<sup>-1</sup> |
| $\beta_h$ | Probability of a human getting infected after an infectious bite | unity |
| $n$ | Number of LLIN campaigns | unity |
| $u_i$ | LLIN usage corresponding to $i^{th}$ campaign, $i \leq i \leq n$ | unity |
| $\tau_i$ | LLIN acquisition time corresponding to $i^{th}$ campaign, $i \leq i \leq n$ | unity |
| $\omega_p$ | Blood feeding inhibition effectiveness of newly acquired LLINs | unity |
| $T_{last}$ | Number of months the data spans | months |
| $\omega_k$ | Insecticidal effectiveness of newly acquired LLINs | unity |
| $T$ | Median survival time of LLINs | months |
| $a$ | Proportion of mosquitoes indoor blood meal taken in-bed | unity |
| $p_1$ | Probability of a pre-infectious human becoming symptomatic infectious after latency | unity |
| $p_2$ | Probability of an asymptomatic human becoming symptomatic due to further infectious bites | unity |
| $\sigma_h$ | Rate of progression of pre-infectious humans to infectious classes | month<sup>-1</sup> |
| $\gamma$ | Per capital recovery rate of a symptomatic infectious human due to treatment with ACT | month<sup>-1</sup> |
| $\mu_h$ | Per capital natural death rate of humans | month<sup>-1</sup> |
| $\delta_h$ | Per capital disease-induced death rate of humans | month<sup>-1</sup> |
| $\Lambda_v$ | Average recruitment rate of female *Anopheles* mosquitoes | mosquitoes × month<sup>-1</sup> |
| $\varepsilon_{vi}$ | mean mosquito indoor blood feeding rate | month<sup>-1</sup> |
| $\varepsilon_{vo}$ | mean mosquito outdoor blood feeding rate | month<sup>-1</sup> |
| $q$ | Proportion of susceptible adult mosquitoes that seek blood meal indoors | unity |
| $\mu_v$ | Per capital natural death rate of mosquitoes | month<sup>-1</sup> |
| $\beta_v$ | Probability of a susceptible mosquito getting infected after an infectious blood meal | unity |
| $\sigma_v$ | Rate of progression of pre-infectious mosquitoes to infectious class | month<sup>-1</sup> |
| $\psi$ | Modification parameter due to reduced level of gametocytes in symptomatic infectious humans | unity |
| $\alpha_{io}$ | Rate at which indoor mosquitoes move outdoor | month<sup>-1</sup> |
| $\alpha_{oi}$ | Rate at which outdoor mosquitoes move indoor | month<sup>-1</sup> |
| $k$ | Number of harmonics in the Fourier series representation of reported malaria cases data | unity |
| $a_j, b_j$ | Amplitudes of cosine and sine terms of the Fourier series representation of reported malaria cases data, $1 \leq j \leq k$ | unity |
| $L$ | Fundamental period for the Fourier series representation of reported malaria cases data | unity |

**Table 8:** Fixed model parameters, values and sources.

| Parameters | Baseline values by States |  |  |  |  | Range | Source |
| --- | --- | --- | --- | --- | --- | --- | --- |
|  | Akwa Ibom | Ebonyi | Kebbi | Oyo | Plateau |  |  |
| $\Lambda_h$ | 6 712.9 | 4 129.676 | 6 456.888 | 10 275.24 | 6 702.487 | - | Derived |
| $p_1$ | 0.17 | 0.17 | 0.17 | 0.17 | 0.17 | [0.02 – 0.27] | Derived |
| $n$ | 2 | 2 | 2 | 2 | 2 | - | NMEP (2022) |
| $u_i, i = 1, 2$ | 0.368, 0.178 | 0.494, 0.481 | 0.376, 0.382 | 0.314, 0.312 | 0.384, 0.267 | - | NMEP (2016, 2022) |
| $\tau_i, i = 1, 2$ | -12, 36 | 0, 48 | 0, 36 | 12, 72 | 0, 60 | - | Derived |
| $\omega_p$ | 0.9 | 0.9 | 0.9 | 0.9 | 0.9 | [0.9 – 1] | WHO (2005) |
| $\omega_k$ | 0.8 | 0.8 | 0.8 | 0.8 | 0.8 | [0.8 – 1] | WHO (2005) |
| $T$ | 39.6 | 39.6 | 63.6 | 38.4 | 63.6 | [12 – 72] | Obi et al. (2020) |
| $a$ | $\frac{2}{3}$ | $\frac{2}{3}$ | $\frac{2}{3}$ | $\frac{2}{3}$ | $\frac{2}{3}$ | $[\frac{1}{4} - \frac{5}{12}]$ | Assumed |
| $\sigma_h$ | 3 | 3 | 3 | 3 | 3 | [2.14 – 3] | Derived |
| $\gamma$ | 2.422 | 3.957 | 3.80 | 3.875 | 3.066 | [2 – 29] | Derived |
| $\mu_h$ | $1.489 \times 10^{-3}$ | $1.521 \times 10^{-3}$ | $1.475 \times 10^{-3}$ | $1.489 \times 10^{-3}$ | $1.673 \times 10^{-3}$ | [0.001 – 0.002] | Derived |
| $\delta_h$ | $7.454 \times 10^{-5}$ | $8.264 \times 10^{-5}$ | $1.24 \times 10^{-5}$ | $9.285 \times 10^{-5}$ | $7.313 \times 10^{-5}$ | - | MAP (2022) |
| $\Lambda_v$ | 298 690 948 | 119 892 200 | 3 888 378 828 | 529 013 373 | 489 819 978 | - | Derived |
| $q$ | 0.357 | 0.639 | 0.466 | 0.451 | 0.531 | [0 – 1] | Derived |
| $\mu_v$ | 2.5 | 2.5 | 2.5 | 2.5 | 2.5 | [2.14 – 4.29] | Derived |
| $\sigma_v$ | 2.143 | 2.143 | 2.143 | 2.143 | 2.143 | [1.67 – 3] | Derived |
| $\psi$ | 0.62 | 0.62 | 0.62 | 0.62 | 0.62 | [0 – 1] | Derived |
| $\varepsilon_{vi}$ | 12.0474 | 10.3539 | 2.1452 | 9.3738 | 21.4178 | [1 – 30] | Derived |
| $\varepsilon_{vo}$ | 3.2392 | 3.6462 | 0.6851 | 2.5264 | 2.3975 | [1 – 30] | Derived |

##### Coefficients of periodic function

To reduce the computational cost of fitting model (16) to data, it is necessary to minimise the number of parameters to be estimated. In pursuance of this, we exclude the coefficients of the periodic function proposed to capture the periodic fluctuation in the observed data of uncomplicated malaria cases from parameters to be fitted. These coefficients will be obtained using the Fourier series representations of the signals, that is, the observed data of uncomplicated malaria cases. Given a time discrete periodic signal *x*[*t*], *t* ∈ ℤ_+_, the Fourier series representation of *x*[*t*] is known to be

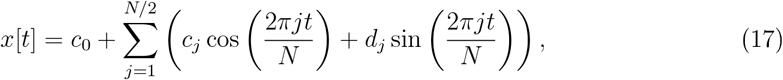

where the coefficients *c*_0_, *c*_*j*_ and *d*_*j*_ represent the average value of *x*[*t*], amplitudes of the cosine and sine terms respectively, and *N* is the length of *x*[*t*]. Factorising *c*_0_, (17) becomes

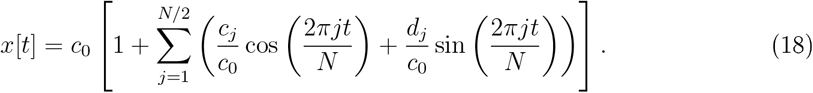

Now, setting *a*_*j*_ = *c*_*j*_*/c*_0_, *b*_*j*_ = *d*_*j*_*/c*_0_, *k* = *N/*2 and *L* = *N*, we have the periodic function proposed to capture the periodic fluctuation in the observed data of uncomplicated malaria cases, that is cases, that is

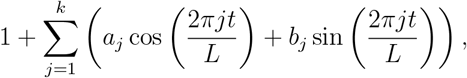

which can be determined explicitly since *c*_0_, *c*_*j*_, *d*_*j*_, *k* and *N* can be obtained, that is *N* = 120, *k* = 60,

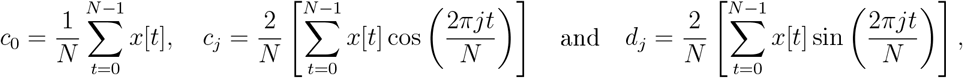

for *j* = 1, 2, · · ·, *k*.

The remaining parameters, *β*_*h*_, *β*_*v*_, *p*_2_, *α*_*io*_ and *α*_*oi*_ will be estimated using available datasets.

##### Bayesian framework for model fitting

Bayesian modelling provides a principled way to quantify uncertainty and incorporate both data and prior knowledge into the model estimates (Gelman et al., 1995; Carpenter et al., 2017; Roda, 2020; Grinsztajn et al., 2021). Stan (CmdStan) is a probabilistic programming framework that supports many probability densities, matrix operations, and numerical ODE solvers primarily used for Bayesian inference (Carpenter et al., 2017; Grinsztajn et al., 2021). In this work, we employed the full Bayesian inference method using Markov Chain Monte Carlo (MCMC), specifically with the dynamic Hamiltonian Monte Carlo (HMC) sampler, which is one of the inference methods that Stan supports and we scripted the Stan programme using the R software.

In Bayesian approach of parameter estimation, the posterior distribution of parameters *θ* given data *y* (the set of plausible parameter values conditional on the data), *p*(*θ*|*y*) is equivalent to the product of prior distribution of the parameters *p*(*θ*) and likelihood of the data given the parameters *p*(*y*|*θ*) (the sampling distribution) (Gelman et al., 1995; Roda, 2020; Grinsztajn et al., 2021). That is,

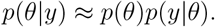

The sampling distribution provides a link between the data and the desired output of the ODEs model to be fitted (Grinsztajn et al., 2021; Roda, 2020). This is done by first determining the distribution of the data and then making the desired ODEs model output a parameter of the set distribution (Grinsztajn et al., 2021; Roda, 2020). Since it is possible to fit different outputs of the ODEs model to different available datasets (Grinsztajn et al., 2021; Roda, 2020), for improved inference, we simultaneously fitted different outputs of the ODEs model to corresponding available datasets.

The sampling distributions linking each dataset and the corresponding ODEs model outputs are hereby described: The 2015 to 2024 monthly reported uncomplicated malaria cases being count data with overdispersion is taken to follow a negative binomial distribution (Bolker, 2008), having the incidence component of the ODEs model, Inc_ODEs_ multiplied by the case reporting probability *r*_*p*_ as its expected value and overdispersion parameter *ϕ*_1_ to be estimated, that is

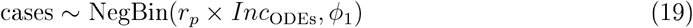

where “cases” is the monthly reported data of uncomplicated malaria cases. The case reporting probability, *r*_*p*_ gives the proportion of the ODEs model-estimated malaria cases that are reported. Other datasets assumed to follow negative binomial distribution are yearly total human population, population of female *Anopheles* mosquitoes indoors and outdoors, with the expected values coming from the ODEs model and overdispersion parameters *ϕ*_2_, *ϕ*_3_ and *ϕ*_4_ respectively, so that

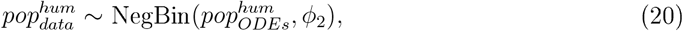

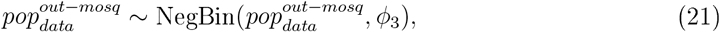

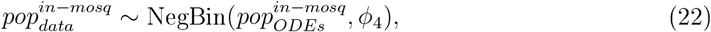

where 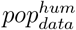 and 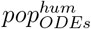 represent data on human population and ODEs model based estimated human population respectively, 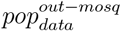 and 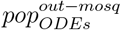 represent data and ODEs based estimated outdoor mosquito population respectively and, 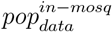 and 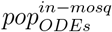 are the data on indoor mosquito population and ODEs based estimated population of indoor mosquitoes respectively. Other datasets used are assumed to follow a binomial distribution as they account for the number of humans or mosquitoes that tested positive to either *P. falciparum* (humans) or sporozoite (mosquitoes) out of certain number tested, hence we have

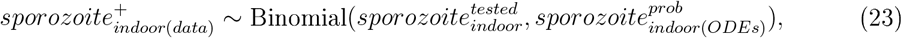

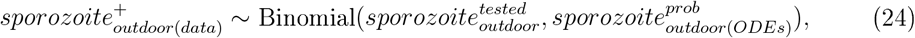

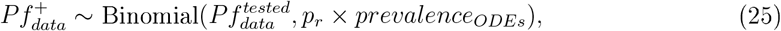

where 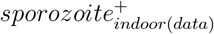 and 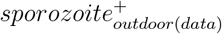 represent the number of mosquitoes that tested positive to sporozoites indoors and outdoors respectively out of the number tested indoor and outdoor 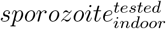 and 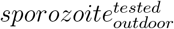 respectively. The ODEs based probability of mosquitoes testing positive to sporozoites indoors and outdoors are respectively 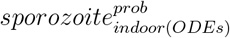 and 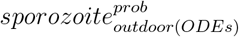. 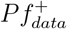 is the number of people who tested positive for *P falciparum* among those tested 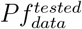, *prevalence*_*ODEs*_ is the ODEs estimated prevalence adjusted with *p*_*r*_, the prevalence reporting probability. Hence, we have the complete list of all sampling distributions used.

Next, to fully describe the Bayesian framework, we give the prior information on the parameters to be estimated (those related to the ODEs and those related to the sampling distributions). Parameters *β*_*h*_, *β*_*v*_, *p*_2_ are constrained within the interval [0,1] and uninformative prior distribution 𝒩 (0, 0.5) was defined to ensure they maintain their meaning as probabilities and proportion. Parameters *α*_*io*_ and *α*_*io*_ are constrained within the interval [0, ∞) and uninformative prior 𝒩 (0, 10) is specified for both of them. Also, parameters *p*_*r*_ and *r*_*p*_ are constrained between interval [0, 1] and uninformative prior distribution *Beta*(1, 2) specified for both of them. Finally, parameters *ϕ*_*i*_ (*i* = 1, 2, 3, 4), each 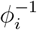 is constrained within the interval [0, ∞) with uninformative prior distribution *Exp*(5) for each 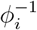.

Due to computational limits, the model was fitted using two parallel MCMC chains with 2000 iterations specified for each chain. While more chains could enhance convergence, convergence diagnostics (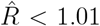 and adequate effective sample sizes) confirmed good mixing and stable estimates. Summaries of estimated parameter and fitted trajectories are shown in Table 9 and Figure 5, respectively.

**Table 9:** Posterior means and 95% credible intervals (CrI) of estimated parameters across states.

| Parameter | Akwa Ibom | Ebonyi | Kebbi | Oyo | Plateau |
| --- | --- | --- | --- | --- | --- |
| $\beta_h$ | 0.97 (0.89–1.00) | 0.97 (0.88–1.00) | 0.98 (0.93–1.00) | 0.75 (0.59–0.94) | 0.98 (0.91–1.00) |
| $\beta_v$ | 0.12 (0.09–0.14) | 0.19 (0.16–0.22) | 0.68 (0.44–0.94) | 0.22 (0.16–0.28) | 0.07 (0.05–0.08) |
| $p_2$ | 0.94 (0.80–1.00) | 0.95 (0.83–1.00) | 0.19 (0.13–0.28) | 0.94 (0.79–1.00) | 0.87 (0.66–1.00) |
| $\alpha_{io}$ | 16.4 (8.1–29.3) | 23.7 (14.6–34.8) | 7.8 (3.2–14.2) | 17.7 (9.4–29.6) | 10.6 (3.7–21.1) |
| $\alpha_{oi}$ | 1.26 (0.38–2.91) | 1.41 (0.74–2.38) | 0.03 (0.00–0.11) | 1.39 (0.51–2.94) | 14.7 (6.1–26.1) |
| $r_p$ | 0.006 (0.006–0.007) | 0.015 (0.014–0.016) | 0.014 (0.013–0.014) | 0.008 (0.007–0.009) | 0.007 (0.007–0.008) |
| $p_r$ | 0.58 (0.50–0.67) | 0.63 (0.55–0.71) | 0.86 (0.79–0.92) | 0.48 (0.42–0.55) | 0.51 (0.44–0.59) |
| $\phi_1^{-1}$ | 0.013 (0.010–0.018) | 0.016 (0.012–0.021) | 0.040 (0.030–0.052) | 0.011 (0.008–0.014) | 0.16 (0.12–0.21) |
| $\phi_2^{-1}$ | 0.003 (0.001–0.011) | 0.021 (0.007–0.061) | 0.040 (0.012–0.13) | 0.014 (0.005–0.040) | 0.020 (0.006–0.060) |
| $\phi_3^{-1}$ | 0.40 (0.19–0.78) | 0.74 (0.38–1.30) | 0.28 (0.07–0.73) | 0.87 (0.47–1.49) | 0.97 (0.16–3.72) |
| $\phi_4^{-1}$ | 1.15 (0.60–1.98) | 1.98 (1.19–2.99) | 0.89 (0.35–1.77) | 1.45 (0.82–2.29) | 3.72 (0.14–3.10) |

**Figure 5.**
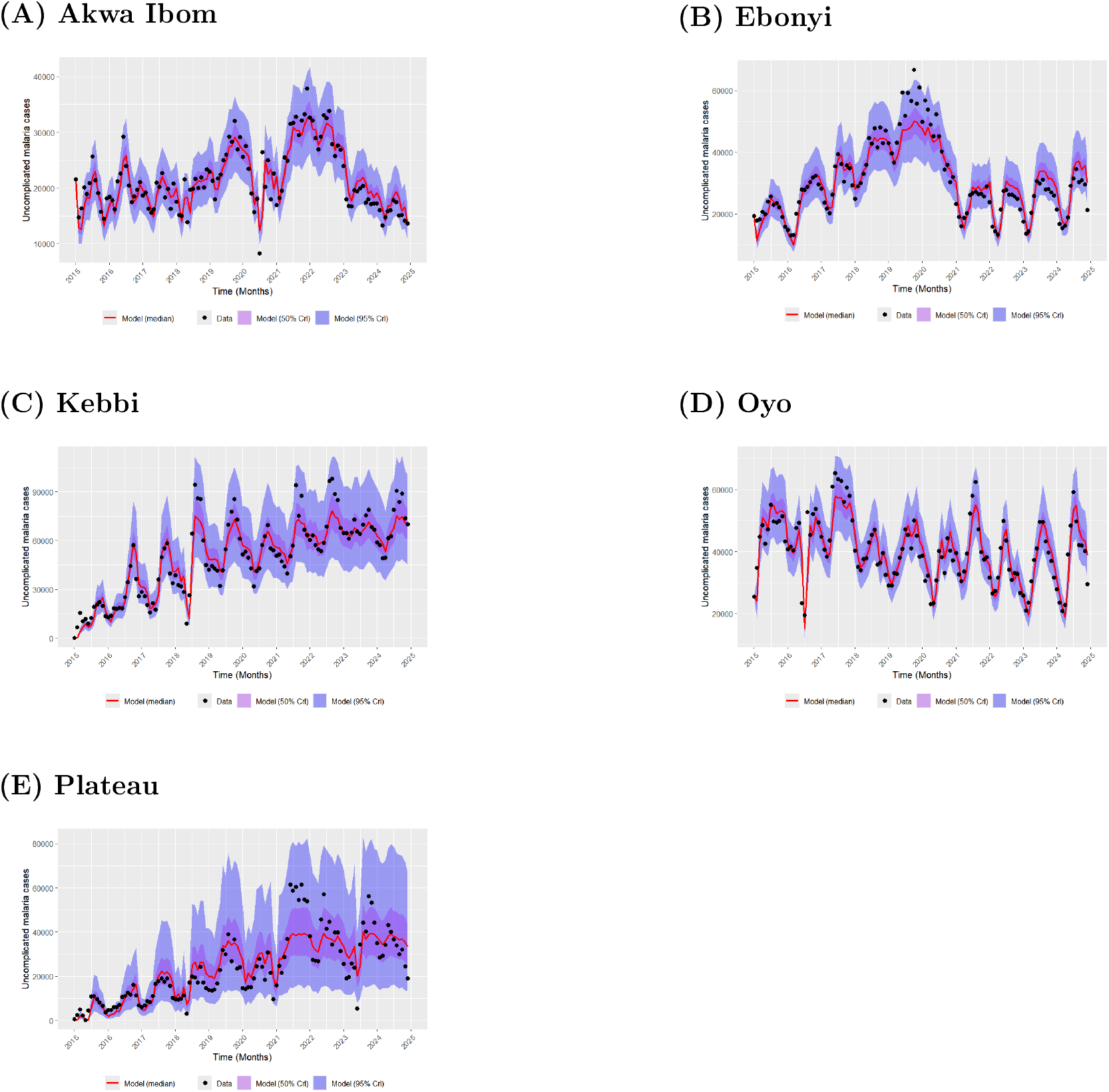
Fitted trajectories of model (16) to monthly uncomplicated malaria case data from Akwa Ibom, Ebonyi, Kebbi, Oyo, and Plateau States.

#### 2.3.2 Goodness of fit

To quantify the goodness of fit of model (16) to reported uncomplicated malaria cases across all states under consideration, we use the normalized index of agreement (NIA) as the metric for goodness of fit. Index of agreement (IA) is a measure of the degree of model prediction error which varies between 0 and 1 (Willmott et al., 1985; Moriasi et al., 2007; Willmott et al., 2012). IA value of 1 indicates a perfect agreement between the observed and fitted values while 0 indicates no agreement (Willmott et al., 1985; Moriasi et al., 2007; Willmott et al., 2012).

Mathematically,

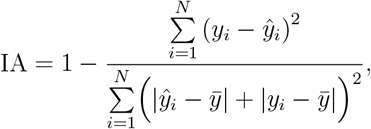

where *y*_*i*_ are the reported values of uncomplicated malaria cases, *ŷ*_*i*_ are the fitted values (posterior medians, that is, 50th percentiles), 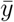 is the mean of the reported values of uncomplicated malaria cases and *N* is the number of observations.

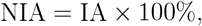

hence NIA measures the percentage of agreement between the reported cases and fitted values. There is more than 80% agreement between the model and the reported values of uncomplicated malaria cases for each state under consideration indicating that the fitting outcomes are good (see Table 10).

**Table 10:** The normalised index of agreement of model (16) to uncomplicated malaria cases.

| State | Akwa Ibom | Ebonyi | Kebbi | Oyo | Plateau |
| --- | --- | --- | --- | --- | --- |
| NIA (%) | 94.62 | 96.09 | 97.01 | 93.57 | 92.61 |

#### 2.3.3 Scenario analysis

Using the stand alone generated quantity in CmdStanR, posterior distribution of the estimated parameters are used for further simulations (Stan Development Team, 2024). With 2015 to 2024 time frame, we simulated counterfactual scenarios in which no outdoor biting activities of female *anopheles* mosquitoes were recorded, there was no gap between LLIN ownership and actual usage and LLIN replacement was carried out in three years time interval.

Percentage of households with at least one net reported in NMEP (2016) and NMEP (2022) was used to represent LLIN ownership achieved through the same LLIN campaigns that led to LLIN usage *u*_1_ and *u*_2_ in Table 8. Replacing the values of *u*_1_ and *u*_2_ for each state with LLIN ownership to simulate the counterfactual scenario in which there was no gap between LLIN ownership and actual usage, we have *u*_1_ = 74.2% and *u*_2_ = 41.3% for Akwa Ibom, *u*_1_ = 88.4% and *u*_2_ = 66.2% for Ebonyi, *u*_1_ = 86.7% and *u*_2_ = 72.9% for Kebbi, *u*_1_ = 51.3% and *u*_2_ = 53.7% for Oyo, and *u*_1_ = 78.3% and *u*_2_ = 37.8% for Plateau.

Next, we consider a combined counterfactual scenarios in which there was no gap between LLIN ownership and actual usage and LLIN replacement was carried out in three years time interval. To achieve this, we fixed the first time of LLIN acquisition then increase the time step by 36 (three years interval) to obtain the next time of acquisition until the last possible time of acquisition before the length of the data time is exhausted. Hence, for Akwa Ibom, we have *τ*_1_ = −12, *τ*_2_ = 24, *τ*_3_ = 60 and *τ*_4_ = 96, for Ebonyi *τ*_1_ = 0, *τ*_2_ = 36, *τ*_3_ = 72 and *τ*_4_ = 108, for Kebbi *τ*_1_ = 0, *τ*_2_ = 36, *τ*_3_ = 72 and *τ*_4_ = 108, for Oyo *τ*_1_ = 12, *τ*_2_ = 48 and *τ*_3_ = 84 and for Plateau *τ*_1_ = 0, *τ*_2_ = 36, *τ*_3_ = 72 and *τ*_4_ = 108. Since originally, there were no LLIN campaign at the new time points assumed, to generate the hypothetical LLIN ownership associated with these new time points, we fixed the first LLIN ownership for each state to correspond to the first time of LLINs acquisition, and assume the same LLIN ownership for the next hypothetical time point until we reach the second real time of acquisition. For any hypothetical time point up to and above second real time, we assign the LLIN ownership of the second real time of acquisition till we exhaust the length of the data time. Hence, LLIN usage corresponding to the times of acquisition in the three year replacement plan are: *u*_1_ = 74.2%, *u*_2_ = 74.2%, *u*_3_ = 41.3% and *u*_4_ = 41.3% for Akwa Ibom; *u*_1_ = 88.4%, *u*_2_ = 88.4%, *u*_3_ = 66.2% and *u*_4_ = 66.2% for Ebonyi; *u*_1_ = 86.7%, *u*_2_ = 72.9%, *u*_3_ = 72.9% and *u*_4_ = 72.9% for Kebbi; *u*_1_ = 51.3%, *u*_2_ = 51.3% and *u*_3_ = 53.7% for Oyo; and *u*_1_ = 78.3%, *u*_2_ = 78.3%, *u*_3_ = 37.8% and *u*_4_ = 37.8% for Plateau.

We further estimated the number of cases during the period between 2015 and 2024 that were attributable to the outdoor biting activities of female *anopheles* mosquitoes, gap between LLIN ownership and actual usage, as well as non-compliance to the three years replacement strategy of LLINs recommended by WHO. This is achieved by estimating the number of cases that would have been if those factors were absent and then taking the difference between the number of cases produced by this situation and the number of cases in the baseline where the factors were actually present.

Lastly, we assume a three year replacement strategy for LLINs and sweep through different levels of LLIN usage between 0 and 1 as well as different levels of percentage reduction in the baseline outdoor biting rate between 0 to 100%. We simulated the total malaria cases with all combinations of LLIN usage and outdoor biting rates obtained to determine how LLIN usage and outdoor biting activities of mosquitoes interdependently affect the outcome of malaria cases.

## 3 Results

Here, we present the results of the simulated scenarios. Figures 6 present the trajectories of malaria cases under the counterfactual scenario of no gap between LLIN ownership and usage compared with the baseline scenario (i.e., the current situation) for all selected states. To keep the plots simple, only posterior median trajectories are shown, while the 95% credible intervals are omitted.

**Figure 6.**
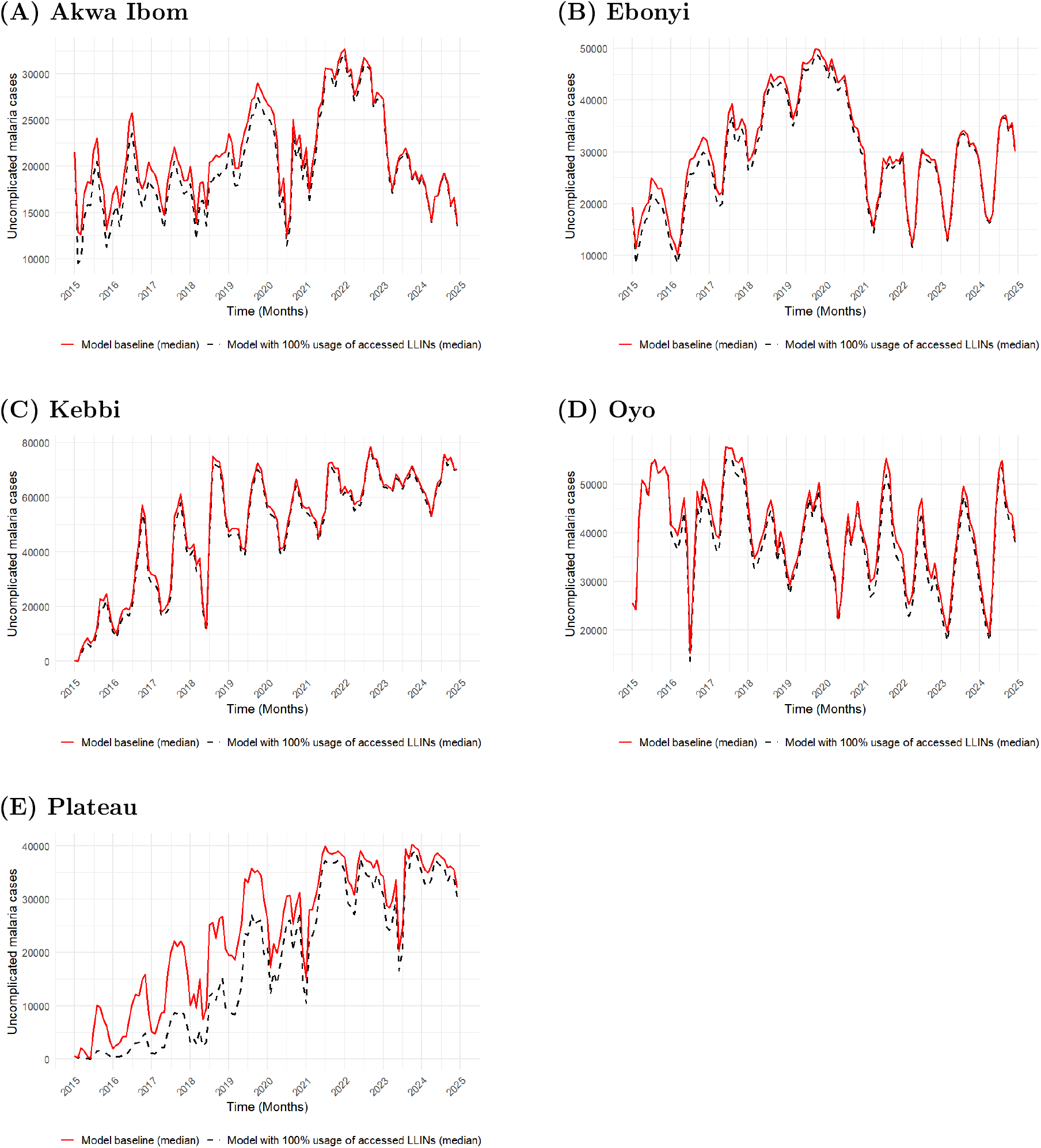
Plots showing simulated scenarios of 100% usage of owned LLINs compared with model baseline for Akwa Ibom, Ebonyi, Kebbi, Oyo and Plateau States.

In Akwa Ibom (Figure 6(A)), replacing actual usage levels (*u*_1_ = 36.8%, *u*_2_ = 17.8%) with ownership levels (74.2%, 41.3% at *τ*_1_ = −12 |2014] and *τ*_2_ = 36 |2018]) results in only a marginal reduction in cases, which largely disappears by mid-2022. Similarly, in Ebonyi (Figure 6(B)), using all owned LLINs (88.4% in 2015 and 66.2% in 2019) produces only a modest deviation from baseline usage (49.4%, 48.1%). For Kebbi (Figure 6(C)), the trajectories under full usage of owned nets (86.7%, 72.9% in 2015 and 2018) and the baseline (37.6%, 49.4%) are nearly indistinguishable. In Oyo (Figure 6(D)), applying full usage of owned nets (51.3%, 53.7% in 2016 and 2021) produces only slight improvements over actual usage (31.4%, 31.2%). Finally, for Plateau (Figure 6(E)), the trajectories under full usage (78.3%, 37.8% in 2015 and 2020) and baseline usage (38.4%, 26.7%) are the most distinct compared to what is observed for other states.

Figure 7 presents plots of simulated scenario in which there is 100% usage of all LLINs accessed and three year LLIN replacement strategy is adhered to across all selected states. Figure 7(A) shows reduced malaria cases for the counterfactual scenario than the baseline throughout for Akwa Ibom. One difference observable in this scenario away from what was seen in the scenario where all accessed LLINs were used only is that the trajectory of the counterfactual scenario are kept clearly away from the baseline throughout. Indeed, this is expected as the LLINs are replenished before becoming ineffective unlike the previous scenario where the LLINs were not replaced on time. Similarly, Figure 7(B) also shows a bit of further reduction in malaria cases for Ebonyi with the counterfactual scenario. For Kebbi, Figure 7(C) shows that the counterfactual scenario still struggles to be distinct from the baseline though there is a slight difference from the previous scenario. However, the case of Oyo is similar to what was observed for Akwa Ibom and Ebonyi having further reduction in malaria cases as seen in Figure 7(D). Plateau again shows further reduction that is more pronounced than what is observed in other states as seen in Figure 7(E). It should be noted that the latter and the former scenarios are simulated in the presence of the baseline outdoor biting rate.

**Figure 7.**
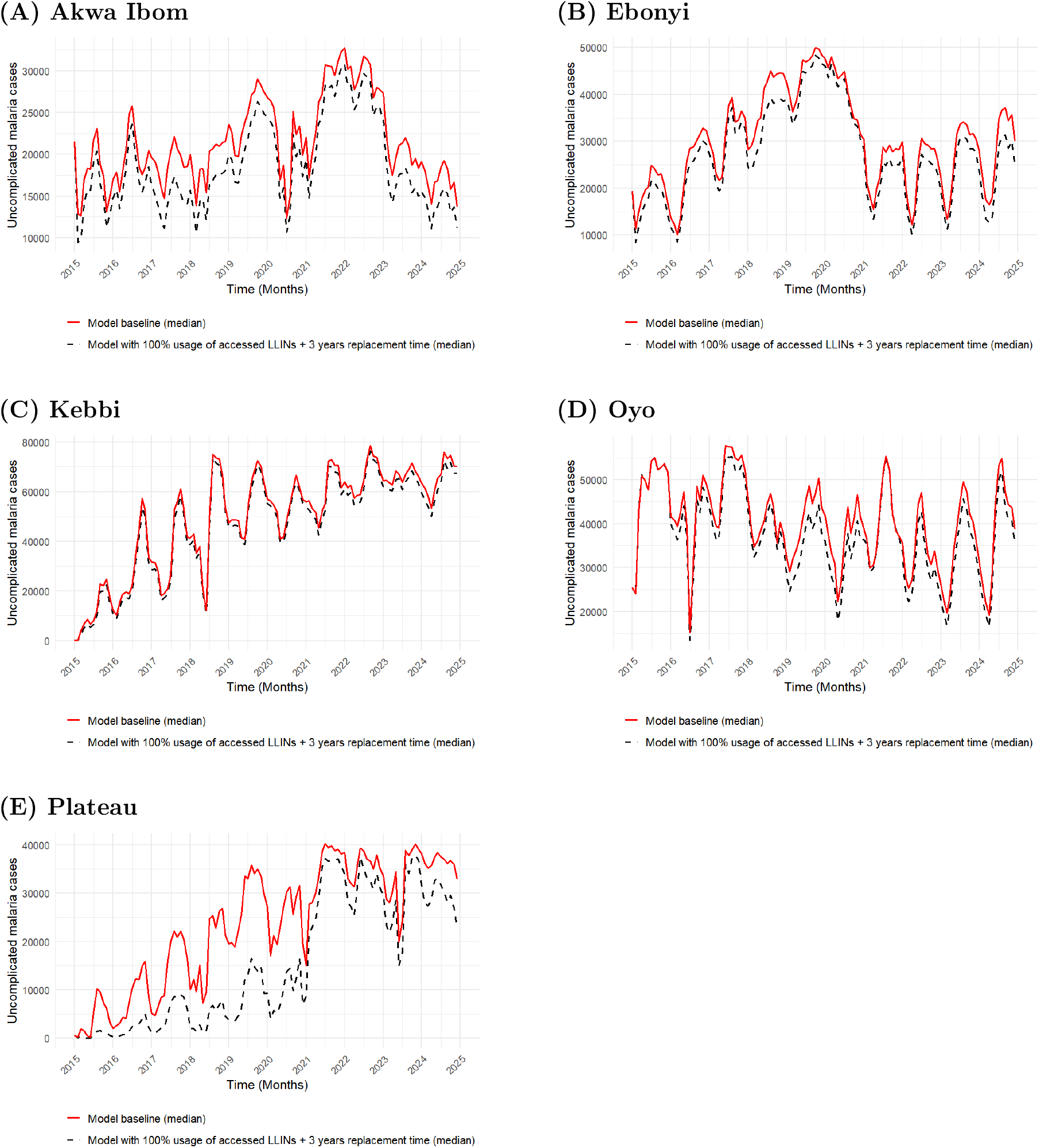
Plots showing scenarios with 100% usage of owned LLINs and 3 years replacement time compared with model baseline for Akwa Ibom, Ebonyi, Kebbi, Oyo and Plateau State.

Plots of the scenario with no outdoor biting are presented in Figure 8 for all selected states. For Akwa Ibom, Figure 8(A) shows a huge decline in the number malaria cases over time with the maximum being less than 10 000 cases. This sharp decline is common to Ebonyi (Figure 8(B)), Kebbi (Figure 8(C)) and Oyo (Figure 8(D)) except for Plateau (Figure 8(E)) where the scenario with no outdoor biting and the baseline are almost indistinguishable.

**Figure 8.**
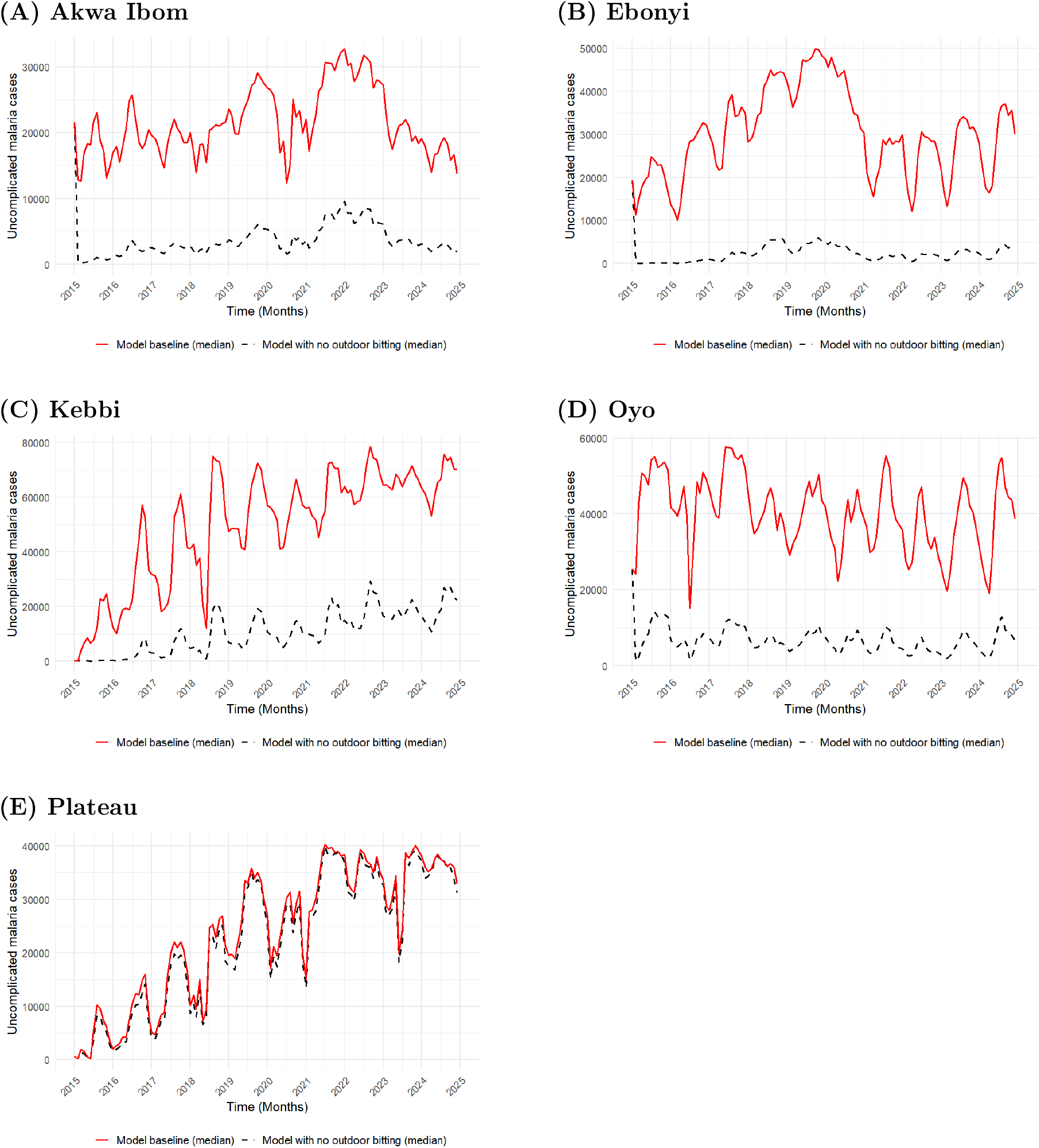
Plots showing simulated scenarios with no outdoor biting compared with model baseline for Akwa Ibom, Ebonyi, Kebbi, Oyo and Plateau States.

Next, we present the estimated total number of malaria cases under the simulated scenarios over the entire study time (2015-2025) across all selected states; additional cases under the baseline scenario attributable to behavioural gap between baseline and counterfactual scenarios estimated by taking difference of total cases in the baseline and total cases in a given counter-factual scenario; and the percentage of these additional cases in the baseline scenario that are attributable to behavioural gap between baseline and counterfactual scenarios.

In Akwa Ibom, Ebonyi, Kebbi and Oyo states, the counterfactual scenario of eliminating outdoor biting produces far greater reductions in malaria cases compared to other behavioural scenarios. For instance, in Akwa Ibom (Table 11), achieving 100% usage of accessed LLINs reduces cases by only 162, 228 (95% CrI 153, 593−171, 127), representing a 6.26% (95% CrI 5.79− 6.80) decline. When timely three-year replacement is added, the reduction nearly doubles to 349, 317 (95% CrI 308, 196−401, 287), equivalent to 13.49% (95% CrI 11.51−15.95). In contrast, eliminating outdoor biting achieves a much larger difference of 2, 135, 146 (95% CrI 1, 974, 751− 2, 254, 170), corresponding to an 82.44% (95% CrI 73.97 − 89.58) decline. A similar pattern is observed in Ebonyi and Oyo, while in Kebbi the gains from moving beyond eliminating the own-ership—usage gap to also including three-year replacement are modest. Howbeit, the situation in Plateau state exhibits a different pattern, where the counterfactual scenario of eliminating outdoor biting produces the least reduction in malaria cases compared to other behavioural scenarios as shown in the table.

**Table 11:** Estimated total and attributable malaria cases under baseline and counterfactual behavioural scenarios in five Nigerian states, with posterior mean and 95% credible intervals (CrI)

| State | Scenario | Total estimated cases (95% CrI) | Attr. cases (95% CrI) | Attr. % (95% CrI) |
| --- | --- | --- | --- | --- |
| Akwa Ibom | Baseline | 2,591,833<br>(2,515,704–2,669,494) | – | – |
|  | UG | 2,429,605<br>(2,344,987–2,513,043) | 162,228<br>(153,593–171,127) | 6.26 (5.79–6.80) |
|  | TR | 2,242,516<br>(2,114,416–2,364,437) | 349,317<br>(308,196–401,287) | 13.49 (11.51–15.95) |
|  | OB | 456,687<br>(262,225–694,744) | 2,135,146<br>(1,974,751–2,254,170) | 82.44 (73.97–89.58) |
| Ebonyi | Baseline | 3,652,134<br>(3,536,222–3,774,603) | – | – |
|  | UG | 3,476,697<br>(3,356,267–3,600,975) | 175,437<br>(171,818–179,986) | 4.81 (4.63–5.04) |
|  | TR | 3,283,111<br>(3,130,556–3,427,021) | 369,023<br>(347,846–405,079) | 10.11 (9.21–11.46) |
|  | OB | 292,349<br>(160,792–470,425) | 3,359,785<br>(3,302,898–3,378,120) | 92.03 (87.54–95.45) |
| Kebbi | Baseline | 5,997,312<br>(5,673,300–6,329,061) | – | – |
|  | UG | 5,782,374<br>(5,452,694–6,122,536) | 214,938<br>(203,955–225,265) | 3.59 (3.26–3.90) |
|  | TR | 5,698,820<br>(5,321,319–6,069,672) | 298,493<br>(268,191–355,218) | 4.99 (4.26– 6.26) |
|  | OB | 1,435,517<br>(419,586–3,103,499) | 4,561,795<br>(3,224,731–5,250,319) | 76.37 (50.96–92.60) |
| Oyo | Baseline | 4,858,124 (4,732,824 – 4,996,739) | – | – |
|  | UG | 4,622,908<br>(4,481,026–4,757,744) | 235,216<br>(228,480–249,743) | 4.84 (4.62–5.26) |
|  | TR | 4,503,237<br>(4,334,187–4,659,546) | 354,887<br>(332,885–396,368) | 7.31 (6.68–8.39) |
|  | OB | 826,400<br>(491,683–1,254,625) | 4,031,724<br>(3,742,253–4,239,899) | 83.04 (74.89–89.61) |
| Plateau | Baseline | 3,002,845<br>(2,685,876–3,368,055) | – | – |
|  | UG | 2,308,573<br>(2,050,501–2,598,847) | 694,272<br>(640,287–768,409) | 23.14 (22.52–23.83) |
|  | TR | 1,909,258<br>(1,675,713–2,167,036) | 1,093,586<br>(1,012,828–1,201,301) | 36.45 (35.51–37.62) |
|  | OB | 2,829,458<br>(2,519,965–3,204,028) | 173,387<br>(160,491–181,925) | 5.79 (4.84–6.26) |

UG = No LLIN usage gap; TR = Timely LLIN replacement and No LLIN usage gap; OB = Outdoor biting eliminated; Attr. cases = Baseline – Counterfactual; Attr. % = (Baseline – Counterfactual)/Baseline × 100.

Figure 9 summarizes the potential percentage reduction in malaria cases achievable by independently addressing human and vector behavioural factors in each state, inferred as the proportion of cases attributable to these behaviours.

**Figure 9.**
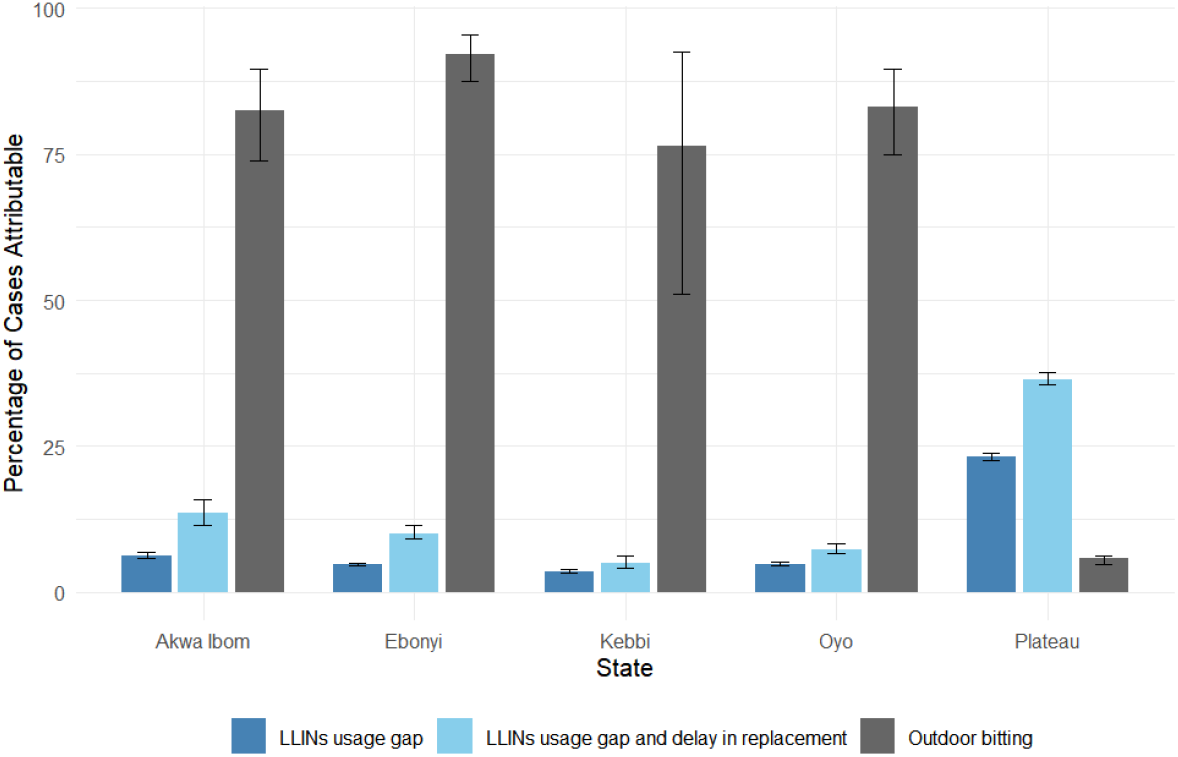
Bar plots showing the potential percentage reduction in malaria cases (with error bars representing 95% crI) achievable by independently targetting human and vector behavioural factors across different states in Nigeria.

Figure 10 shows how total malaria cases respond to different combinations of LLIN usage and reductions in baseline outdoor biting rates, assuming a three-year LLIN replacement cycle across the five study states. Case burden is represented by a colour gradient ranging from purple (lowest) to yellow (highest). Overall, results from Akwa Ibom, Ebonyi, Oyo, and Kebbi indicate that higher LLIN usage substantially reduces malaria cases when outdoor biting is low; however, this protective effect weakens as outdoor biting increases.

**Figure 10.**
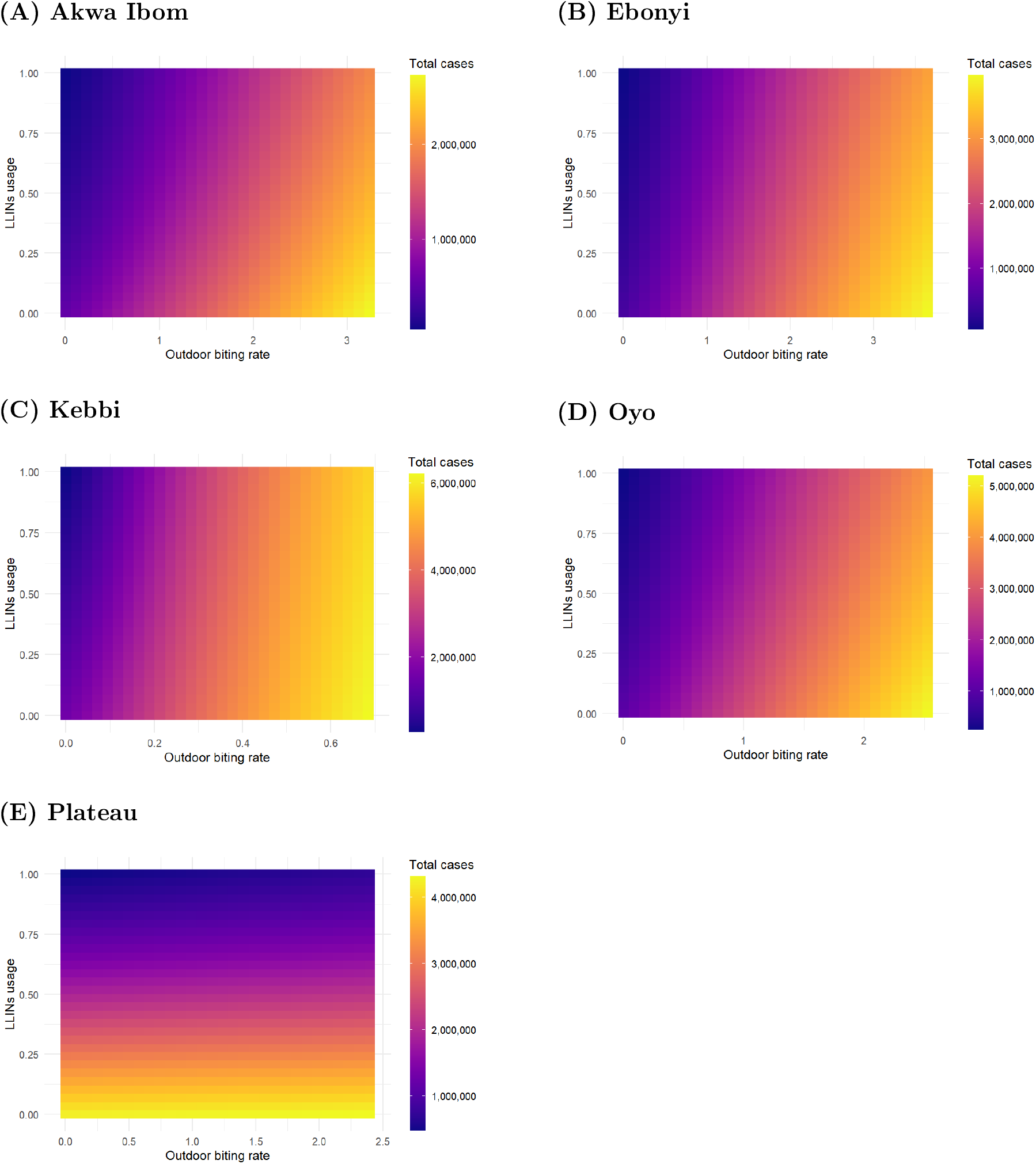
Plots showing variation in total malaria cases over a range of percentage LLIN usage and percentage reduction in baseline outdoor biting rate in Akwa Ibom, Ebonyi, Kebbi, Oyo and Plateau States.

In Akwa Ibom (Figure 10(A)), total malaria cases approach three million, with LLINs remaining highly effective when outdoor biting rates are within 0—1 bites per mosquito per month—corresponding to at least a 69% reduction in the baseline biting rate. Beyond this range, LLIN effectiveness declines even with increased usage. A similar pattern is observed in Ebonyi (Figure 10(B)), where total cases approach four million, and LLIN effectiveness is maintained only when outdoor biting rates remain between 0—1, equivalent to at least a 73% reduction in baseline biting.

In Kebbi (Figure 10(C)), total cases exceed six million, with LLINs effective only when outdoor biting remains below 0.2—representing a reduction of at least 71% in baseline biting. For Oyo (Figure 10(D)), total cases slightly exceed five million, and LLIN protection remains effective up to about 0.8 bites per mosquito per month, corresponding to a 68% reduction in baseline outdoor biting.

By contrast, in Plateau (Figure 10(E)), total cases exceed four million, and increasing LLIN usage consistently enhances its protective effect across all outdoor biting rates.

## 4 Discussion

The use of long-lasting insecticidal nets (LLINs) remains one of the most effective and widely implemented interventions for malaria prevention. However, their effectiveness is constrained by behavioural and operational challenges such as non-usage, delayed replacement of worn-out nets, and their limited protection against mosquitoes that bite outdoors (Monroe et al., 2020). Nonetheless, the mere existence of these challenges does not necessarily imply equal epidemiological significance; their relative impacts must be quantified. For instance, Bradley et al. (2015) found that although malaria vectors in Bioko Island, Equatorial Guinea exhibited outdoor biting behaviour that could potentially undermine control efforts, this factor did not significantly drive malaria transmission.

Recognising that these limiting factors vary across ecological and social settings in Nigeria, this study quantified their relative contributions to persistent malaria transmission. Using counterfactual analysis, we estimated the potential reduction in malaria incidence that could be achieved if each constraint—outdoor mosquito biting, LLIN usage gaps, and delayed LLIN replacement—were eliminated.

Our results indicate that eliminating outdoor mosquito biting would have produced substantial reductions in malaria burden across Akwa Ibom, Ebonyi, Kebbi, and Oyo states over the past decade (2015—2024). Specifically, estimated reductions were: Akwa Ibom — 82.4% (95%, CrI : 74.0 − 89.6); Ebonyi — 92.0% (95%, CrI : 87.5 − 95.5); Kebbi — 76.4% (95%, CrI : 51.0 − 92.6); and Oyo — 83.0% (95%, CrI : 74.9 − 89.6). These findings clearly identify outdoor biting as a major driver of sustained malaria transmission in these states.

In contrast, closing the gap between LLIN ownership and actual usage alone produced relatively modest reductions: Akwa Ibom — 6.3%, Ebonyi — 4.8%, Kebbi — 3.6%, and Oyo — 4.8%. When both usage gaps and delayed replacement were addressed simultaneously, the combined effect remained limited, with reductions ranging from 5% to 13% across states. These results suggest that human behavioural factors such as LLIN usage gap and non-adherence to replacement schedules have a smaller impact on sustained transmission compared to outdoor mosquito activity.

Importantly, the sharp contrast between the effects of eliminating outdoor biting and imroving LLIN usage should not be interpreted as evidence that LLINs are unimportant. Rather, outdoor biting likely acts as an effect modifier, weakening the relationship between LLIN use and malaria incidence reduction. The parameter sweep analysis supports this interpretation, showing that LLIN protective effectiveness reduces as outdoor biting increases. While higher LLIN usage consistently reduces transmission, its benefits are limited in areas with intense outdoor mosquito biting activity. Furthermore, the threshold beyond which outdoor biting undermines LLIN effectiveness varies across states, likely reflecting ecological differences in the abundance and infectivity of outdoor-biting mosquito populations.

The implications of these findings are far-reaching. Exposure to outdoor biting emerged as a major contributor to ongoing malaria transmission in Akwa Ibom, Ebonyi, Kebbi, and Oyo states. This likely reflects a behavioural adaptation among local mosquito populations—shifting towards exophagic (outdoor-biting) and exophilic (outdoor-resting) behaviours—to evade contact with insecticide-treated nets. Additionally, cultural and occupational patterns such as evening social gatherings, agricultural work, and informal night-time trading increase human exposure to outdoor bites. Other factors may include proximity of mosquito breeding sites to human residential areas. From a policy standpoint, these findings underscore the urgent need to expand malaria control beyond indoor-based interventions. Specifically, the NMEP and state malaria programmes should adopt integrated vector management (IVM) strategies combining environmental management, larval source control, and continued LLIN promotion to address both indoor and outdoor transmission dynamics.

In Plateau State, however, outdoor biting contributed the least to sustained transmission (5.8%, 95%, CrI : 4.8 − 6.3), while the larger contributor was the gap between LLIN ownership and usage (23.1%, 95%, CrI : 22.5−23.8). When combined with delays in replacement, the total contribution rose to 36.5% (95%, CrI : 35.5 − 37.6). This suggests that in Plateau, behavioural factors—particularly low LLIN use—play a more prominent role than vector behaviour. Hence, substantial reductions in malaria incidence could be achieved by increasing LLIN usage and ensuring adherence to the three-year replacement cycle, even under current outdoor biting conditions. From a policy perspective, this calls for behavioural change communication and community-level engagement to promote consistent LLIN use. Evidence suggests that low usage is commonly driven by discomfort from heat, fear of suffocation, net damage, poor maintenance, and misconceptions about malaria transmission (Monroe et al., 2020; NMEP, 2022). To address these barriers, the NMEP and Plateau State Ministry of Health should: Implement targeted public awareness campaigns using culturally appropriate channels to promote LLIN usage; Strengthen net replacement logistics to maintain the three-year cycle; and Engage local health workers and community leaders to improve net adoption and accountability.

Overall, the differing contributions of human and vector behavioural factors across states underscore the need for state-specific surveillance to monitor local entomological and behavioural dynamics. Such information is critical for tailoring malaria control strategies to local contexts and maximizing intervention effectiveness

Our findings are consistent with previous studies showing that increased LLIN use and timely replacement reduce malaria transmission (Killeen et al., 2006; Agusto et al., 2013; Ngonghala, 2023), while also extending this understanding by highlighting the modifying influence of outdoor biting (Sherrard-Smith et al., 2019). Specifically, Sherrard-Smith et al. (2019) reported that across Africa, increased outdoor transmission could lead to approximately 10.6 million additional malaria cases each year, even under universal LLIN and IRS coverage. Their findings further highlighted that outdoor biting substantially reduces the number of malaria cases averted through vector control interventions.

As previously noted by Ngonghala et al. (2014), assuming a fixed LLIN effectiveness may overestimate its true impact. Our results similarly indicate that neglecting outdoor biting in transmission models can lead to overestimation, as it implies that LLINs protect against all mosquito biting behaviour—a simplification not supported by empirical data. This limitation is evident in many models that assess LLIN effectiveness, which often assume both constant net efficacy and homogeneous mosquito biting behaviour.

While, it is well established that understanding human and vector behavioural factors in a local setting is critical for selecting appropriate vector control intervention (Khatib et al., 2025)—Our study further highlights the importance of estimating the actual impact of these factors on malaria burden. Such estimates are essential for making evidence-based decisions on which interventions to prioritise.

Methodologically, this study refined the representation of LLIN biological activity and durability presented in Ngonghala et al. (2014) and Ngonghala (2023). Based on field evidence that LLIN effectiveness varies across ecological settings and net types (WHO, 2005; Randriamaherijaona et al., 2017; Obi et al., 2020; Hiruy et al., 2023), LLIN protective and mosquito-killing effectiveness were parametrised using empirically reported median survival times rather than a uniform three-year assumption for lost of insecticidal properties. Moreover, recognizing that LLIN chemical efficacy often remains optimal throughout the net surviving time (Obi et al., 2020; Hiruy et al., 2023), mosquito-killing effectiveness was modelled to scale with net survivability rather than assuming a declining chemical efficacy. Finally, incorporating the actual timing of LLIN mass distribution campaigns instead of adopting a hypothetical replacement cycle further improved alignment between simulated interventions and real-world implementation, enhancing the reliability of state-specific transmission estimates.

## 5 Conclusions

This study quantified the unaverted malaria cases in Nigeria over the past decade (2015—2024) attributable to three key factors: outdoor mosquito biting, the gap between LLIN ownership and usage, and delays in LLIN replacement. The goal was to determine which of these factors most strongly sustains malaria transmission and to provide evidence to guide strategic priorities for the NMEP. To achieve this, we developed a deterministic compartmental model (*S*_*h*_*E*_*h*_*A*_*h*_*I*_*h*_*S*_*h*_—*S*_*vi*_*E*_*vi*_*I*_*vi*_—*S*_*vo*_*E*_*vo*_*I*_*vo*_) that captures malaria transmission between humans and both indoor- and outdoor-biting *Anopheles* mosquito populations. The model accounts for the waning protective and killing effectiveness of LLINs over time and was calibrated using monthly malaria case data, supported by demographic, entomological, and prevalence information to improve parameter robustness. Model fitting showed strong agreement with observed data, and counterfactual simulations were performed to isolate and quantify the contributions of the identified behavioural and entomological factors.

Our results reveal that outdoor mosquito biting is the dominant contributor to sustained malaria transmission in Akwa Ibom, Ebonyi, Kebbi, and Oyo states, while LLIN usage gaps and delayed replacement play comparatively minor roles. Conversely, Plateau State exhibits the opposite pattern, where low LLIN usage and irregular replacement cycles are the primary behavioural drivers of continued transmission, with outdoor biting contributing only marginally.

These findings have significant implications for malaria control policy in Nigeria. They demonstrate that while increasing LLIN coverage and ensuring timely replacement remain essential, these efforts alone will not achieve optimal reductions in malaria incidence in states where outdoor mosquito biting is widespread. Integrating LLIN-based interventions with strategies that specifically target outdoor transmission—such as environmental management, spatial repellents, larval source control, and improved housing designs—will be critical for achieving long-term malaria control in such settings. In contrast, in Plateau State, behavioural change interventions that encourage consistent LLIN use and adherence to the three-year replacement cycle should be prioritized to maximize existing control efforts.

This study is not without limitations. First, it assumes that LLIN acquisition occurred simultaneously across the population, whereas in reality, people acquire nets at different time. Second, the model uses average indoor and outdoor biting rates, preventing assessment of seasonal variations in their effects on malaria incidence. Third, the Fourier forcing function applied is restricted to the calibration interval, limiting predictions beyond 2024 (Karami et al., 2025). Finally, the study did not explicitly incorporate complementary interventions such as environmental management, which could influence outdoor mosquito abundance and, consequently, malaria transmission. Future research should therefore aim to integrate these additional factors and extend model predictions beyond the calibration period using more flexible forcing frameworks.

Despite these limitations, this retrospective analysis provides actionable insights to strengthen Nigeria’s malaria control programme. By quantifying the contributions of human and vector behavioural factors to persistent malaria transmission, the study offers evidence-based guidance to help the NMEP prioritize interventions.

In conclusion, while maintaining high LLIN coverage, usage, and timely replacement remains fundamental, eliminating outdoor biting exposure is crucial to achieving further reductions in malaria burden. A shift toward integrated, state-specific vector management strategies—addressing both human behaviour and vector adaptation—is essential for achieving and sustaining malaria elimination goals in Nigeria.

## Data Availability

All data produced in the present study are available upon reasonable request to the authors

https://nmdrnigeria.ng/dhis-web-commons/security/login.action

## CRediT authorship contribution statement

**Conceptualization:** Emmanuel A. Bakare, Idowu I. Olasupo. **Data curation:** Emmanuel A. Bakare, Idowu I. Olasupo. **Formal analysis:** Idowu I. Olasupo, Emmanuel A. Bakare. **Funding acquisition:** Emmanuel A. Bakare. **Investigation:** Idowu I. Olasupo, Emmanuel A. Bakare. **Methodology:** Idowu I. Olasupo, Emmanuel A. Bakare. **Software:** Idowu I. Olasupo, Emmanuel A. Bakare. **Supervision:** Emmanuel A. Bakare, L. O. Salaudeen. **Writing** — **original draft:** Idowu I. Olasupo. **Writing — review & editing:** Emmanuel A. Bakare, Idowu I. Olasupo.

## Acknowledgments

The authors are grateful to the ICAMMDA team, WAMCAD, NMEP, and Malaria Consortium for their resourceful support.

## Code and data availability

The R codes used to generate the results presented in this manuscript are available from the corresponding author upon a reasonable request. The dataset used is not publicly available; however, access can be gained upon reasonable request from the NMEP through https://nmdrnigeria.ng/dhis-web-commons/security/login.action.

## References

Abebaw, A., Aschale, Y., Kebede, T., and Hailu, A. (2022). The prevalence of symptomatic and asymptomatic malaria and its associated factors in debre elias district communities, northwest ethiopia. Malaria Journal, 21(1):167.

Afolabi, B., Amajoh, C., Adewole, T., and Salako, L. (2006). Seasonal and temporal variations in the population and biting habit of mosquitoes on the atlantic coast of lagos, nigeria. Medical Principles and Practice, 15(3):200–208.

Agusto, F. B., Del Valle, S. Y., Blayneh, K. W., Ngonghala, C. N., Goncalves, M. J., Li, N., Zhao, IL. and Gong, H. (2013). The impact of bed-net use on malaria prevalence. Journal of theoretical biology, 320:58–65.

Bakare, E. (2015). On the qualitative behaviour of a human-mosquito model for malaria with multiple vector control strategies. International Journal of Ecological Economics and Statistics, 36(1):96–113.

Bolker, B. M. (2008). Ecological models and data in R. Princeton University Press.

Bradley, J., Lines, J., Fuseini, G., Schwabe, C., Monti, F., Slotman, M., Vargas, D., Garcia, G., Hergott, D., and Kleinschmidt, I. (2015). Outdoor biting by anopheles mosquitoes on bioko island does not currently impact on malaria control. Malaria Journal, 14(1):170.

Carpenter, B., Gelman, A., Hoffman, M. D., Lee, D., Goodrich, B., Betancourt, M., Brubaker, M. A., Guo, J., Li, P., and Riddell, A. (2017). Stan: A probabilistic programming language. Journal of statistical software, 76.

GDC (2024). CDC - DPDx - Malaria, https://www.cdc.gov/dpdx/malaria/index.html. [Accessed 07-09-2024],

Chitnis, N., Hyman, J. M., and Cushing, J. M. (2008). Determining important parameters in the spread of malaria through the sensitivity analysis of a mathematical model. Bulletin of mathematical biology, 70:1272–1296.

Collins, O. and Duffy, K. (2022). A mathematical model for the dynamics and control of malaria in nigeria. Infectious Disease Modelling, 7(4):728–741.

Davis, E. L., Hollingsworth, T. D., and Keeling, M. J. (2020). A novel age-structured mosquito model for assessing the mechanisms behind vector control success.

Davis, E. L., Hollingsworth, T. D., and Keeling, M. J. (2024). An analytically tractable, age-structured model of the impact of vector control on mosquito-transmitted infections. PLOS Computational Biology, 20(3):el011440.

Degefa, T., Githeko, A. K., Lee, M.-C., Yan, G., and Yewhalaw, D. (2021). Patterns of human exposure to early evening and outdoor biting mosquitoes and residual malaria transmission in ethiopia. Acta tropica, 216:105837.

Fernandez Montoya, L., Alafo, C., Martí-Soler, H., Máquina, M., Comiche, K., Cuamba, L, Munguambe, K., Cator, L., Aide, P., Galatas, B., et al. (2022). Overlaying human and mosquito behavioral data to estimate residual exposure to host-seeking mosquitoes and the protection of bednets in a malaria elimination setting where indoor residual spraying and nets were deployed together. Plos one, 17(9):e0270882.

Gelman, A., Carlin, J. B., Stern, H. S., and Rubin, D. B. (1995). Bayesian data analysis second edition corrected version (30 jan 2008).

Gimba, B. and Bala, S. I. (2017). Modeling the impact of bed-net use and treatment on malaria transmission dynamics. International Scholarly Research Notices, 2017(1):6182492.

Grinsztajn, L., Semenova, E., Margossian, C. C., and Riou, J. (2021). Bayesian workflow for disease transmission modeling in stan. Statistics in medicine, 40(27):6209–6234.

Hiruy, II. N., Irish, S. R., Abdelmenan, S., Wuletaw, Y., Zewde, A., Woyessa, A., Haile, M., Chibsa, S., Lorenz, L., Worku, A., et al. (2023). Durability of long-lasting insecticidal nets (llins) in ethiopia. Malaria Journal, 22(1):109.

Irikannu, K., Onyido, A., Nwankwo, E., Umeanaeto, P., Onwube, O., Ogaraku, J., Ezeagwuna, D., Onyebueke, A., and Okoduwa, A. (2020). A survey of man-biting mosquito species in a tropical rainforest community in the south eastern nigeria. Environment and Ecology, 38(3):290–299.

Karami, H., Chowell, G., Mujica, O. J., and Smirnova, A. (2025). Parameter Estimation and Forecasting Strategies for Cholera Dynamics: Insights from the 1991-1997 Peruvian Epidemic. Mathematics, 13(10):1692.

Khatib, B., Mcha, J., Pandu, Z., Haji, M., Hassan, M., Ali, H., Mrisho, R., Abdallah, K., Ali, A., Ali, K., Said, T., Mohamed, S., Mkali, H., Mgata, S., Makwaruzi, S., Gulaka, M., Makenga, G., Mkude, S., Githu, V., Mero, V., Serbantez, N., Ballard, S.-B., Chan, A., Shija, S. J., and Govella, N. J. (2025). Early evening outdoor biting by malaria-infected anopheles arabiensis vectors threatens malaria elimination efforts in Zanzibar. Malaria Journal, 24.

Killeen, G. F., Kihonda, J., Lyimo, E., Oketch, F. IL. Kotas, M. E., Mathenge, E., Schellenberg, J. A., Lengeler, C., Smith, T. A., and Drakeley, C. J. (2006). Quantifying behavioural interactions between humans and mosquitoes: evaluating the protective efficacy of insecticidal nets against malaria transmission in rural tanzania. BMC infectious diseases, 6(1):1–10.

Lindblade, K. A., Steinhardt, L., Samuels, A., Kachur, S. P., and Slutsker, L. (2013). The silent threat: asymptomatic parasitemia and malaria transmission. Expert review of anti-infective therapy, 11(6):623–639.

Mandala, W. L., Harawa, V., Dzinjalamala, F., and Tembo, D. (2021). The role of different components of the immune system against plasmodium falciparum malaria: Possible contribution towards malaria vaccine development. Molecular and biochemical parasitology, 246:111425.

MAP (2022). Malaria Atlas Project | Data, https://data.malariaatlas.org/trends?year=2022&metricGroup=Malaria&geographicLevel=adminl&metricSubcategory=Pf&metricType=rate&metricName=incidence. [Accessed 12-09-2024],

Milali, M. P., Sikulu-Lord, M. T., and Govella, N. J. (2017). Bites before and after bedtime can carry a high risk of human malaria infection. Malaria journal, 16(1):1–10.

Monroe, A., Moore, S., Okumu, F., Kiware, S., Lobo, N. F., Koenker, H., Sherrard-Smith, E., Gimnig, J., and Killeen, G. F. (2020). Methods and indicators for measuring patterns of human exposure to malaria vectors. Malaria Journal, 19.

Moriasi, D. N., Arnold, J. G., Van Liew, M. W., Bingner, R. L., Harmel, R. D., and Veith, T. L. (2007). Model evaluation guidelines for systematic quantification of accuracy in watershed simulations. Transactions of the ASA BE. 50(3):885–900.

Mosha, J. F., Lukole, E., Charlwood, J. D., Wright, A., Rowland, M., Bullock, O., Manjurano, A., Kisinza, W., Mosha, F. W., Kleinschmidt, I., et al. (2020). Risk factors for malaria infection prevalence and household vector density between mass distribution campaigns of long-lasting insecticidal nets in north-western tanzania. Malaria journal, 19:1–11.

Moshi, I. R., Manderson, L., Ngowo, H. S., Mlacha, Y. P., Okumu, F. O., and Mnyone, L. L. (2018). Outdoor malaria transmission risks and social life: a qualitative study in southeastern tanzania. Malaria journal, 17(1):1–11.

Ngonghala, C. N. (2023). The impact of temperature and decay in insecticide-treated net efficacy on malaria prevalence and control. Mathematical Biosciences, 355:108936.

Ngonghala, C. N., Del Valle, S. Y., Zhao, R., and Mohammed-Awel, J. (2014). Quantifying the impact of decay in bed-net efficacy on malaria transmission. Journal of theoretical biology, 363:247–261.

NMEP (2016). Nigeria Malaria Indicator Survey 2015 Final Report. Abuja, Nigeria, and Rockville, Maryland, USA: NMEP, NPC, and IGF.

NMEP (2021). National Malaria Strategic Plan, 2021-2025. Federal Ministry of Health, Abuja, Nigeria.

NMEP (2022). Nigeria Malaria Indicator Survey, 2021. Federal Ministry of Health, Abuja, Nigeria.

NMEP, LSHTM, and KEMRI-Wellcome Trust Research Programme (2018). Nigeria: A Profile of Malaria Control and Epidemiology, 2018 Overview. Federal Ministry of Health, Abuja, Nigeria and the Department for International Development, UK.

NMEP and NPC and ICF (2022). Nigeria Malaria Indicator Survey 2021 Final Report. Abuja, Nigeria, and Rockville, Maryland, USA: NMEP, NPC, and ICF.

NPC (2024). Nigeria population projections and demographic indicators. https://nationalpopulation.gov.ng/publications. [Accessed 25-08-2024],

Obi, E., Okoh, F., Blaufuss, S., Olapeju, B., Akilah, J., Okoko, O. O., Okechukwu, A., Maire, M., Popoola, K., Yahaya, M. A., et al. (2020). Monitoring the physical and insecticidal durability of the long-lasting insecticidal net dawaplus® 2.0 in three states in Nigeria. Malaria journal, 19:1–19.

Ojurongbe, O., Lawal, O. A., Abiodun, O. O., Okeniyi, J. A., Oyeniyi, A. J., and Oyelami, O. A. (2013). Efficacy of artemisinin combination therapy for the treatment of uncomplicated falciparum malaria in nigérian children. The journal of infection in developing Countries, 7(12):975–982.

Ordinioha, B. (2012). The use and misuse of mass distributed free insecticide-treated bed nets in a semi-urban community in rivers state, nigeria. Annals of African medicine, 11(3):163–168.

Oyewole, I., Awolola, T., Ibidapo, C., Oduola, A., Okwa, O., and Obansa, J. (2007). Behaviour and population dynamics of the major anopheline vectors in a malaria endemic area in southern nigeria. Journal of vector borne diseases, 44(1):56.

PMI (2015). AIRS Nigeria Final Entomology Report. November 2014 - December 2015. Africa Indoor Residual Spraying Project, Abt Associates Inc.

PMI (2016). AIRS Nigeria Final Entomology Report. February - December 2016. Africa Indoor Residual Spraying Project, Abt Associates Inc.

PMI (2017). AIRS Nigeria Final Entomology Report. January — December 2017. Rockville, Maryland, USA: Africa Indoor Residual Spraying Project, Abt Associates Inc.

PMI (2018). The PMI VectorLink Nigeria Project Annual Entomology Report. April — September 2018. The PMI VectorLink Project. Rockville, MD. VectorLink, Abt Associates Inc.

PMI (2019). The PMI VectorLink Nigeria Project Annual Entomology Report. November 2018 September 2019. The PMI VectorLink Project. Rockville, MD. VectorLink, Abt Associates Inc.

PMI (2020). The PMI VectorLink Nigeria Project Annual Entomology Report. October 2019 September 2020. The PMI VectorLink Project. Rockville, MD. VectorLink, Abt Associates Inc.

PMI (2021). The PMI VectorLink Nigeria Project Annual Entomology Report. October 2020 September 2021. The PMI VectorLink Project. Rockville, MD. VectorLink, Abt Associates Inc.

PMI (2022). The PMI VectorLink Nigeria Project Annual Entomology Report. October 2021 September 2022. The PMI VectorLink Project. Rockville, MD. VectorLink, Abt Associates Inc.

PMI (2024). Nigeria Malaria Operational Plan FY 2024. Retrieved from https://www.pmi.gov.

Randriamaherijaona, S., Raharinjatovo, J., and Boyer, S. (2017). Durability monitoring of long-lasting insecticidal (mosquito) nets (llins) in madagascar: physical integrity and insecticidal activity. Parasites & vectors, 10:1–11.

Roda, W. C. (2020). Bayesian inference for dynamical systems. Infectious Disease Modelling, 5:221–232.

Savi, M. K. (2022). An overview of malaria transmission mechanisms, control, and modeling. Medical Sciences, 11(1):3.

Sherrard-Smith, E., Skarp, J. E., Beale, A. D., Fornadel, C., Norris, L. C., Moore, S. J., Mihreteab, S., Charlwood, J. D., Bhatt, S., Winskill, P., Griffin, J. T., and Churcher, T. S. (2019). Mosquito feeding behavior and how it influences residual malaria transmission across africa. Proc. Natl. Acad. Sci. U. S. A., 116(30):15086–15095.

Stan Development Team (2024). Stan User’s Guide Version 2.35.

Tomas, T., Eligo, N., Tamiru, G., and Massebo, F. (2022). Outdoor and early hour human biting activities of malaria mosquitoes and the suitability of clay pot for outdoor resting mosquito collection in malaria endemic villages of southern rift valley, ethiopia. Parasite Epidemiology and Control, 19:e00278.

USAIDs (2019). Malaria operational plan fy 2019. President’s Malaria Initiative Nigeria.

WHO (2005). Guidelines for laboratory and field testing of long-lasting insecticidal mosquito nets. World Health Organization, https://iris.who.int/handle/10665/69007.

WHO (2012). Handbook for integrated vector management. World Health Organization.

WHO (2015). Global technical strategy for malaria 2016-2030. World Health Organization.

WHO (2022). World malaria report 2022. World Health Organization.

WHO (2024). World malaria report 2024.

Willmott, C. J., Ackleson, S. G., Davis, R. E., Feddema, J. J., Klink, K. M., Legates, D. R., O’donnell, J., and Rowe, C. M. (1985). Statistics for the evaluation and comparison of models. Journal of Geophysical Research: Oceans, 90(C5):8995–9005.

Willmott, C. J., Robeson, S. M., and Matsuura, K. (2012). A refined index of model performance. International Journal of climatology, 32(13):2088–2094.

Yakob, L. and Yan, G. (2009). Modeling the effects of integrating larval habitat source reduction and insecticide treated nets for malaria control. PLoS One, 4(9):e6921.

Zemene, E., Belay, D. B., Tiruneh, A., Lee, M.-C., Yewhalaw, D., and Yan, G. (2021). Malaria vector dynamics and utilization of insecticide-treated nets in low-transmission setting in southwest ethiopia: implications for residual transmission. BMC Infectious Diseases, 21(1):882.

